# Predicting the Peak and COVID-19 trend in six high incidence countries: A study based on Modified SEIRD model

**DOI:** 10.1101/2020.12.14.20248117

**Authors:** Punam Bedi, Shivani Dhiman, Neha Gupta, Vinita Jindal, Pushkar Gole

## Abstract

The novel Coronavirus (COVID-19) has claimed the lives of almost a million people across the globe and this trend continues to rise rapidly day by day. The fear of getting infected by Corona virus is affecting the people emotionally, psychologically and mentally. They are not able to work to their full capacity and are also worried about the well beings of their near and dear ones. The National governments have taken up several measures like lockdowns, closing of educational institutions, and work from home for employees of companies wherever feasible. Governments are also advising people to take precautions like not to go out if not necessary, use of mask and keep a distance of appx. 6 ft. if you need to go out as the virus spreads from human to human in close proximity. These measures have helped to limit the spread of this virus in the past few months. However, due to rapid increase in the daily confirmed cases, it is becoming tougher for the governments to provide facilities like testing kits, hospitalization facilities, oxygen cylinders etc. to the infected persons. Thus, there is an urgent need to accurately estimate the number of cases in coming future that can help governments in acquiring the required resources. Further, to handle the economic distress caused by this virus, long-term planning is equally important. Focusing on these two aspects, this paper proposes to use the Modified SEIRD (Susceptible-Exposed-Infected-Recovered-Deceased) model to predict the peak and spread trend of COVID-19 in six countries namely USA, India, Brazil, Russia, Peru and Colombia having the highest number of confirmed cases. As in COVID-19, even infected asymptomatic persons can spread the infection, the chosen model is well suited as exposed compartment of SEIRD model includes asymptomatic exposed individuals which are infectious. Epidemiological data till 9^th^ September 2020 has been utilised to perform short-term predictions till 31^st^ December 2020. Long-term predictions have been computed till 31^st^ December 2023, to estimate the end of the virus in the above-mentioned six nations. Small values of MAPE (Mean Absolute Percentage Error) have been obtained for the models fitted to reported data for all the countries. Student t-test has been used for accepting the predictions of the Modified SEIRD model based on the reported data.

## 1. Introduction

In the second last month of year 2019, a novel coronavirus appeared in the Wuhan city of China (Ma, 2020). From there, it has spread to almost the entire world in a short span of time. It started as a deadly disease, then turned into an epidemic, and later it was declared as a pandemic by the World Health Organization (WHO) on 11^th^ March 2020 (World Health Organization, 2020). The novel coronavirus, also known as Corona Virus Disease 2019 (COVID-19), has affected 215 countries with more than 28 million cases and 0.91 million deaths worldwide as on 9^th^ September 2020. The exponential spread of the virus has uniformly affected the healthcare system of different countries. This can be verified by the fact that the list of worst-affected countries by COVID-19 includes both developed and developing nations.

COVID-19 infection is a respiratory disease that mainly affects the lungs of a patient. Although this infection impacts everyone equally, patients with existing respiratory diseases become easy targets of this infection. Moreover, people with comorbidities also remain at high risk due to their weak immunity. Towards the beginning of this pandemic, it was thought that only elderly population is vulnerable to this virus. However, as time passed, many new cases were reported among children and adults as well. As on 9^th^ September 2020, USA, India, Brazil, Russia and Peru are the six nations that have seen the maximum number of cases. USA, India, Brazil and Russia have more than 6.5, 4.5, 4.2 and 1 million cases respectively, while the count for Peru remains less than a million. Though India ranks second in the number of cases, it has the least number of total cases per million (3391 cases/million), out of the six worst-affected countries. On the other hand, Peru has the maximum number of total cases per million (21677 cases/million). A similar trend is observed for total number of deaths per million with Peru having the highest value (922 deaths per million), while India accounting for only 56 deaths per million among the above-mentioned six nations. However, India’s statistics can be a result of the minimum number of tests per million (39915 tests/million) that are being performed in the country.

The COVID-19 infection spreads by directly coming in contact with infected water droplets from an infected individual when he/she coughs or sneezes. Another source of infection is touching any surface that contains infection. To minimize the spread of infection from an infected person, national governments across the world have urged their citizens to maintain social distancing, wear face masks and wash their hands regularly with soap or use alcohol-based sanitizers. Furthermore, some countries also implemented restrictions in the form of lockdowns where people were confined to their homes for several days. During this time, companies allowed its employees to work from home, educational institutions provided online classes, transportation was limited and people were instructed to follow personal hygiene norms in their daily activities.

Apart from necessary precautions, planning for future is another means of efficiently handling the situation. This requires accurate predictions for the growth trend of the infection in near future. These predictions can help policy makers to take timely decisions and plan in advance to lower the increasing burden on the economy and healthcare sector. In recent times, a large number of research articles have been published that focus on the ongoing pandemic. These publications cover a diverse range of topics including the development of COVID-19 vaccine, impact of COVID-19 on patients with comorbidities, the role of a person’s demographics in his susceptibility to the deadly disease and predicting the trend of the pandemic. This paper focuses on the last category and performs short-term and long-term predictions for six worst affected countries namely USA India, Brazil, Russia, Peru and Colombia.

This paper performs predictions for the spread of COVID-19 in six worst-affected countries of the world using Modified SEIRD model. Short-term predictions depict the progress of the virus till 31^st^ December 2020 and can be utilized by the national governments to take immediate measure for handling the pandemic. Whereas, long-term predictions show the pandemic trend for a longer period (in this paper, taken till the end of year 2023) to help the policy-makers for long-term planning. The epidemiological reported data from 22^nd^ January 2020 till 9^th^ September 2020 has been used for modelling and predictions in this paper.

The remaining paper is organized as follows: Section 2 presents the literature review; Section 3 explains the Modified SEIRD model that has been used to perform predictions; Section 4 explains the experimental setup, followed by Section 5 which presents the results of the experimentation. Section 6 concludes the paper.

## 2. Literature Review

In 2019, the COVID-19 pandemic appeared in China and since then, it has spread to the whole world (Rahmandad, Lim, & Sterman, 2020). Every community has contributed to fight this difficult time. Doctors, nurses, cleanliness workers and police department are now referred to as corona warriors due to their devotion to the needs of the society in these trying times. The community of researchers has also played an important role. There are two ways in which researchers have played their part in the fight against COVID-19. First, by predicting the future trend of the pandemic to help in policy making and second, by working towards the development of a vaccine for the novel coronavirus. From the beginning of the pandemic, various researchers have used different modelling techniques to predict the trend of COVID-19 in different regions of world (Giordano, et al., 2020), (Zhao, et al., 2020), (Bhatnagar, 2020) and (Sturniolo, Waites, Colbourn, Manheim, & Panovska-Griffiths, 2020). Some of these works have been discussed in this section.

Anand et al (Anand, Sabarinath, Geetha, & Somanath, 2020) augmented the standard SIR model by incorporating Quarantine and Testing for the prediction of COVID-19 spread in India. In their paper, pre-lockdown and post lockdown conditions were accessed. The parameters of differential equations were calculated for the time period before lockdown and after lockdown. Optimization method, particularly differential evolution optimizer, were used to calculate the parameters of SIR model with Quarantine and Testing. Bhatnagar (Bhatnagar, 2020) proposed a simple mathematical model in which the spread of COVID-19 infection was modelled. The authors observed that as the infection spreads from one person to another, it effectively creates a Geometric Progression by which the total infected individuals can be estimated. Some constraints were introduced in the model including recovery rate of 14 days which further improved their model and gave better results.

Zhou et al (Zhou, et al., 2020) compared three models for short-term prediction as well as peak prediction for eight most affected countries by COVID-19. The three models namely Logistic, SEIR, and adjusted SEIR were used by the authors. Logistic model mathematically shows the dynamic evolution of infected people which is controlled by the growth rate and population capacity. The difference between SEIR and adjusted SEIR is that first one is implemented considering no intervention whereas the second one includes control measures and intervention. For fitting, 10 days of data was used and 7 days prediction was performed.

Taboe et al (Taboe, Salako, Tison, Ngonghala, & Kakaï, 2020) proposed a mathematical model to predict the course of COVID-19 in West Africa. The paper focused on the control measures for the containment of infection spread due to COVID-19. To analyse the effect of control measures such as social distancing, face mask, contact tracing and mass testing, SEIR model was used. Based on the West-Africa data project, baseline parameters for the mathematical model were calculated. The worst-case scenario was depicted when there were no control measures taken by the government. Whereas following the current health measures, it was deduced that the reduction of 67% in daily cases will be seen at the time of peak in West Africa. And with improved control measures it will further reduce the daily cases 76% to 87%.

Chakraborty et al (Chakraborty & Ghosh, 2020) utilised Auto-Regressive Integrated Moving Average (ARIMA) model and Wavelet model to propose a hybrid model. The authors used their proposed model to predict the trend of COVID-19 for five countries namely Canada, France, India, South Korea and United Kingdom. The hybrid model was used to compute short-term predictions for ten days into future. Similarly, Ceylan (Ceylan, 2020) also made use of the ARIMA models for projecting the spread of the pandemic in Europe’s worst-affected countries. Different ARIMA models were constructed by the author based on different parameter values. Using the best model, predictions were performed for France, Italy and Spain.

Roosa et al (Roosa, et al., 2020) performed short-term predictions of COVID-19 for two Chinese provinces namely Guangdong and Zhejiang. In their paper, the authors computed the total confirmed cases using three models viz. Logistic growth model, Richards growth model and a Sub-epidemic wave model. They adopted a bootstrap approach to calculate the uncertainty bounds for the projection of cumulative number of cases. Ahmed (Ahmed, 2020) studied the effect of patient age, gender and geographical location on the pandemic spread. In his work, the Indian population was divided into six groups to study the regional effect of COVID-19 on a patient. Clustering was used by the author to find the similarity between different groups, while Multiple Linear Regression was used to predict the infection source. For each group, different age distributions were studied and their recovery rates were calculated.

Patrikar et al (Patrikar, Poojary, Basannar, & Kunte, 2020) modified the SEIR model in their work to predict the trend of COVID-19 for India. In their model, the authors analysed the effect of social distancing on the pandemic and obtained different graphs for the same. According to the model given by Poonia et al (Poonia & Azad, 2020), it was concluded that social distancing has been effective in controlling the virus spread in India. In their paper, authors performed short-term predictions of COVID-19 for different states of India in worst case scenario. Pandey et al (Pandey, Chaudhary, Gupta, & Pal, 2020) also studied the trend of this disease in India during the initial days of the outbreak. Short-term predictions for two-weeks were performed by the authors using the SEIR model. In their paper, the effect of interventions was also studied on the progress of the infection within the country. Moreover, the authors used the Regression model to predict the number of confirmed and deceased cases in India.

This paper utilises a compartmental epidemiological model, named Modified SEIRD (Susceptible-Exposed-Infected-Recovered-Deceased) model for projecting the spread of COVID-19 in six countries having the highest number of total cases. Short-term predictions till 31^st^ December 2020 have been computed using the Modified SEIRD model. Further, long-term predictions have also been performed for the six countries having the highest number of total infected cases as on 9^th^ September 2020. The next section presents the Modified SEIRD model that has been used for predictions.

## 3. The Modified SEIRD model used for prediction

The SEIR (Susceptible-Exposed-Infectious-Recovered) model is a compartmental model that divides the population into four mutually exclusive compartments. These compartments are named as Susceptible (S), Exposed (E), Infected (I), and Recovered (R). The SEIR model captures the dynamic nature of a pandemic. As the infection progresses, parts of population shift from one compartment to the other (Brauer, Driessche, & Wu, 2008). In the SEIR model, S(t) represents the individuals who have not been infected by the disease on day t, but they are vulnerable to the infection. E(t) represents the individuals who have been exposed to the disease on day t, by coming in contact with an infected person. However, these people are considered asymptomatic as they have not shown any signs of the disease yet. I(t) represents the individuals who have been infected by the disease on day t. Such people act as carriers of infection and can transmit it further. R(t) represents the individuals who have recovered from the infection or succumbed to it on day t. Some authors refer this compartment as the Removed compartment.

In the SEIR model, the transmission rate β regulates the spread of the infection. It signifies the rate at which susceptible people become infected by the disease. *α* is the onset rate with 1/*α* represents the average latent period of the infection; while 1/*γ* represents the average infectious period. The SEIR model considers few assumptions. It is assumed that apart from the deceased patients, there is no entry or departure from the population. Moreover, recovered individuals are assumed to gain immunity and hence, they cannot spread the infection further. It is also assumed that the number of new infection cases are directly proportional to the infected and exposed population.

The SEIR model consists of a single compartment for both recovered and deceased individuals. This model was enhanced to include a separate compartment for the casualties caused by the disease. The resulting model is known as the SEIRD model where D(t) denotes the individuals who have been deceased due to the pandemic on day t. Infected people become Deceased with a rate *μ*.

The SEIRD model assumes that exposed population is non-infectious, whereas it has been seen in COVID-19 cases that asymptomatic individuals are tested positive and are also responsible for spreading the disease. Therefore, the SEIRD model was modified by incorporating asymptomatic individuals in the exposed compartment. This paper utilises the Modified SEIRD model to include asymptomatic cases in exposed population and the same has been shown in Figure 1 below:

**Figure 1:**
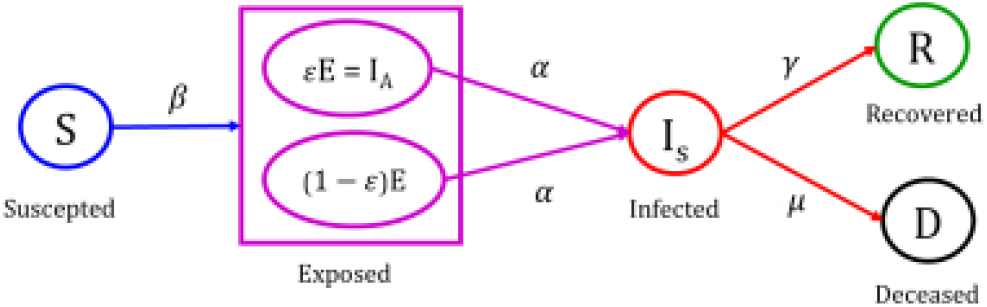
Modified SEIRD model

The Modified SEIRD model has been derived from the SEIRD model. It uses parameter ‘ε’ which represents the Exposed population that is asymptomatic but infectious, and hence infecting others. The µ parameter denotes the rate at which infected people become deceased. The Modified SEIRD model with parameter ε representing asymptotic exposed population is governed by the following differential equations:

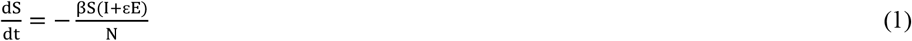

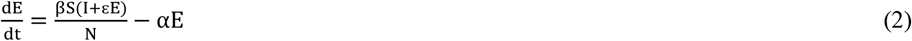

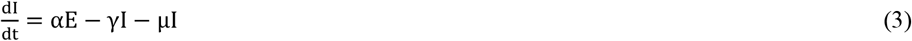

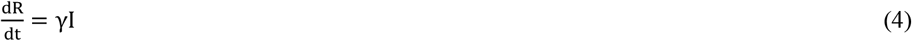

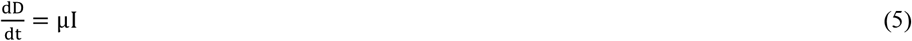

To analyse the pandemic spread, this work utilises the basic reproduction number, Ʀ0. Ʀ0 is the average number of secondary infections generated by an infected person and is defined by equation (6):

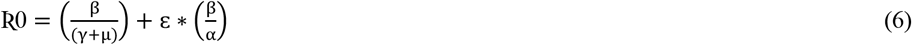

Based on Ʀ0, another parameter named Effective Reproduction number, Ʀe, is defined. Ʀe is the average number of new infections generated by an infectious individual on day, t, and is calculated by equation (7):

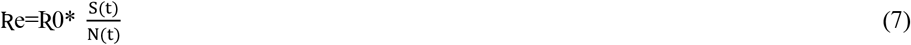

Equations (6) and equation (7) clearly depict the increase by a factor 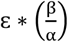 in the value of Ʀ0 and the corresponding increase in Ʀe by the exposed population. Hence the parameter ε, representing the proportion of infectious exposed population, contributes to the growth of COVID-19. High value of this parameter indicates that there are more infectious exposed individuals and contact tracing should be done, so that these individuals can be isolated to reduce the spread of the disease.

## 4. Experimental Setup

In this paper, experiments were performed in Python using Jupyter Notebook. The time-series data of COVID-19 provided by John Hopkins University, was used for experimentation in this paper (Dong, Du, & Gardner, 2020). For the six worst-affected countries, the data of Total Confirmed cases, Total Recovered cases and Total Deceased cases was available since 22^nd^ January 2020. The values of Daily Confirmed cases, Daily Recovered cases and Daily Deceased cases were calculated by subtracting the data values of two consecutive days. Active Infected cases were calculated by subtracting both Total Recovered cases and Total Deceased cases from Total Confirmed cases. The data was pre-processed to remove negative values. Also, the data for each day was replaced with the average of the current data and the data of last four days. This was done to handle untimely reporting of data.

The Modified SEIRD model was used for experimentation, as described in Section 3. The model parameters β, ε, α, γ and µ were optimized by fitting the curve to the reported data. The curve fiiting was performed using the functions provided by Python’s *lmfit* library. For optimization of the above-mentioned parameters, *minimize* function was used with the least square method. The initial value of Active Infected, I0, was taken as 1 assuming the spread started from a single person. The initial values for exposed E0, recovered R0, and deceased D0 were taken as 0. Further, the initial value for Suscpectible S0 was computed using equation (8):

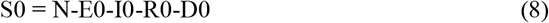

where, N is total population of the place in consideration.

Fitting of data with single set of paramters (β, ε, α, γ and µ) is not appropriate for performing predictions as the reported data is dynamic in nature. Therefore, the curve was fitted to data by computing the optimized parameters (β, ε, α, γ and µ) on a weekly basis which were used for further predictions. To obtain the optimized parameters, the initial values and the bounds for the parameters were given as inputs to the *minimize* function of *lmfit* library. For parameter β, initial value was taken as 0.5 with bounds [0.001,1]. For ε, 0.1 was used as the initial value with [0.0001,1] as bounds; 1/5 was the initial value for α and [1/6,1] were the bounds. For parameter γ, the initial value was 1/10 with bounds were chosen to be [1/150, 1]; and for µ, 0.01 and [0.0001,1] was the initial value and its bounds respectively. The results obtained from the Modified SEIRD model have been presented in the next section.

## 5. Results

This section presents the predictions obtained using Modified SEIRD model. The results for both short-term and long-term predictions are discussed in the next subsections.

### 5.1. Results for Short-Term Predictions

The short-term predictions for the six worst-affected nations, were performed till 31^st^ December 2020. This includes the predictions for seven time-series data namely, Daily Confirmed cases, Daily Recovered cases, Daily Deceased cases, Total Confirmed cases, Total Recovered cases, Total Deceased cases and Active Infected cases. To analyse the results obtained by the model, Case Fatality Rate (CFR), Recovery Rate (RR) and Mean Absolute Percentage Error (MAPE) values have been calculated using equations (9), (10) and (11) respectively.

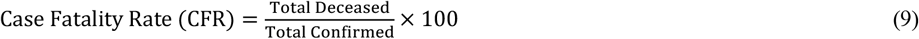

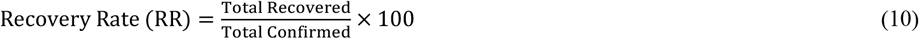

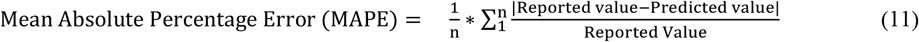

The RR and CFR values for six worst-affected countries on 9^th^ September 2020 (date till the reported data was used for experimentation) have been shown in Table 1 and for short-term predictions, RR and CFR values on 31^st^ December 2020, have been given in Table 2.

**Table 1:** RR and CFR for six worst-affected countries on 9^th^ September 2020

| Country | Reported RR (%) | Predicted RR (%) | Reported CFR (%) | Predicted CFR (%) |
| --- | --- | --- | --- | --- |
| USA | 37.53 | 37.44 | 3.0003 | 3.0004 |
| India | 77.74 | 77.53 | 1.68 | 1.70 |
| Brazil | 86.03 | 85.48 | 3.06 | 3.05 |
| Russia | 82.31 | 82.13 | 1.74 | 1.73 |
| Peru | 77.13 | 75.57 | 4.33 | 4.32 |
| Colombia | 80.50 | 78.53 | 3.21 | 3.22 |

**Table 2:**
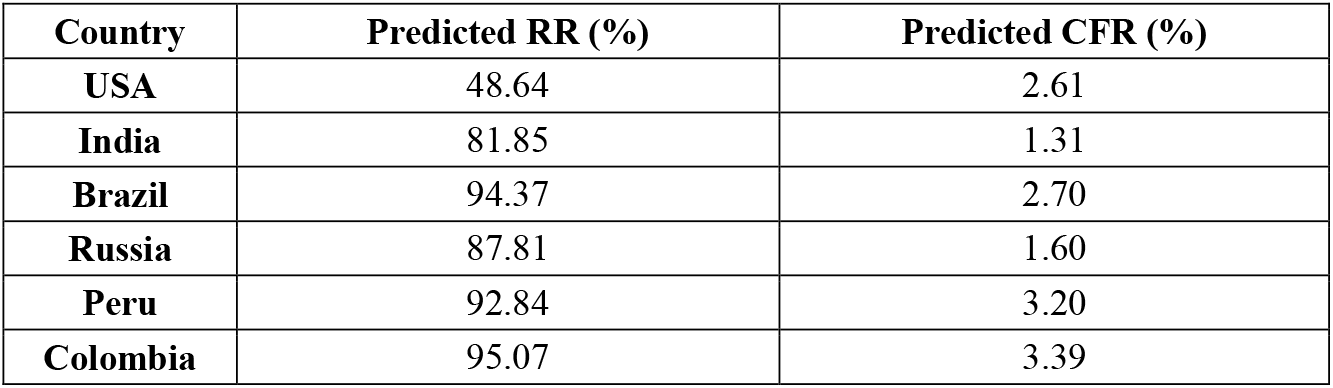
RR and CFR for six worst-affected countries on 31^st^ December 2020

| Country | Predicted RR (%) | Predicted CFR (%) |
| --- | --- | --- |
| USA | 48.64 | 2.61 |
| India | 81.85 | 1.31 |
| Brazil | 94.37 | 2.70 |
| Russia | 87.81 | 1.60 |
| Peru | 92.84 | 3.20 |
| Colombia | 95.07 | 3.39 |

It can be observed from Table 1 that the predicted values of CFR and RR are significantly similar to the corresponding values for reported data on 9^th^ September 2020. It was found that USA has the lowest RR of 37.53 on 9^th^ September 2020, with CFR value as 3.0003. However, by the end of year 2020, it is predicted that this value will increase to 48.64 and its CFR value will decrease to 2.61 as shown in Table 2. On the other hand, Peru has the highest CFR of 4.33, while its RR stands at 77.13. According to the Modified SEIRD model, Peru will achieve the second highest RR of 92.84 by the end of year 2020, among the six countries in consideration. At the same time, Russia will witness a 5% increase in RR and a 0.13% decrease in the value of CFR. Next, Figure 2 (a-f) depict the short-term predictions of Daily Confirmed cases for USA, India, Brazil, Russia, Peru and Colombia respectively.

**Figure 2(a-f):**
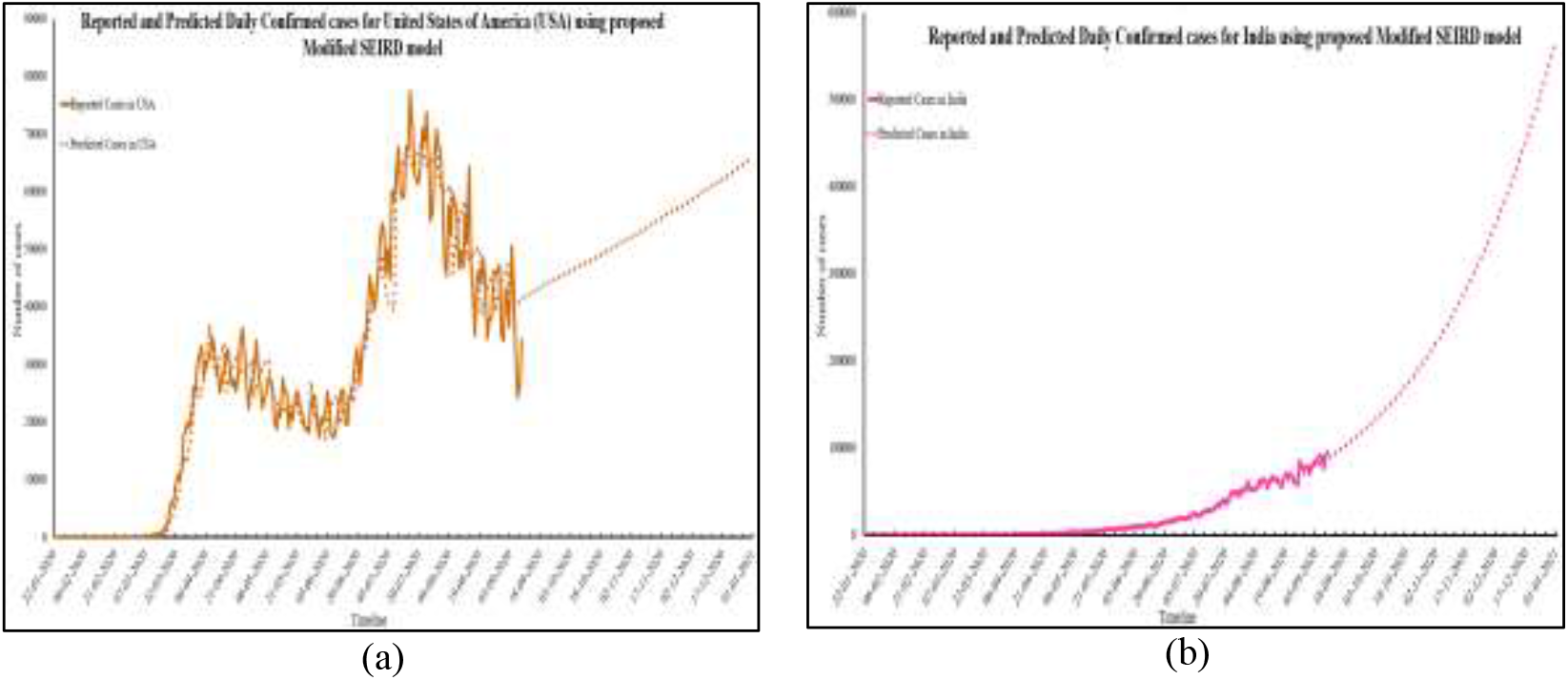
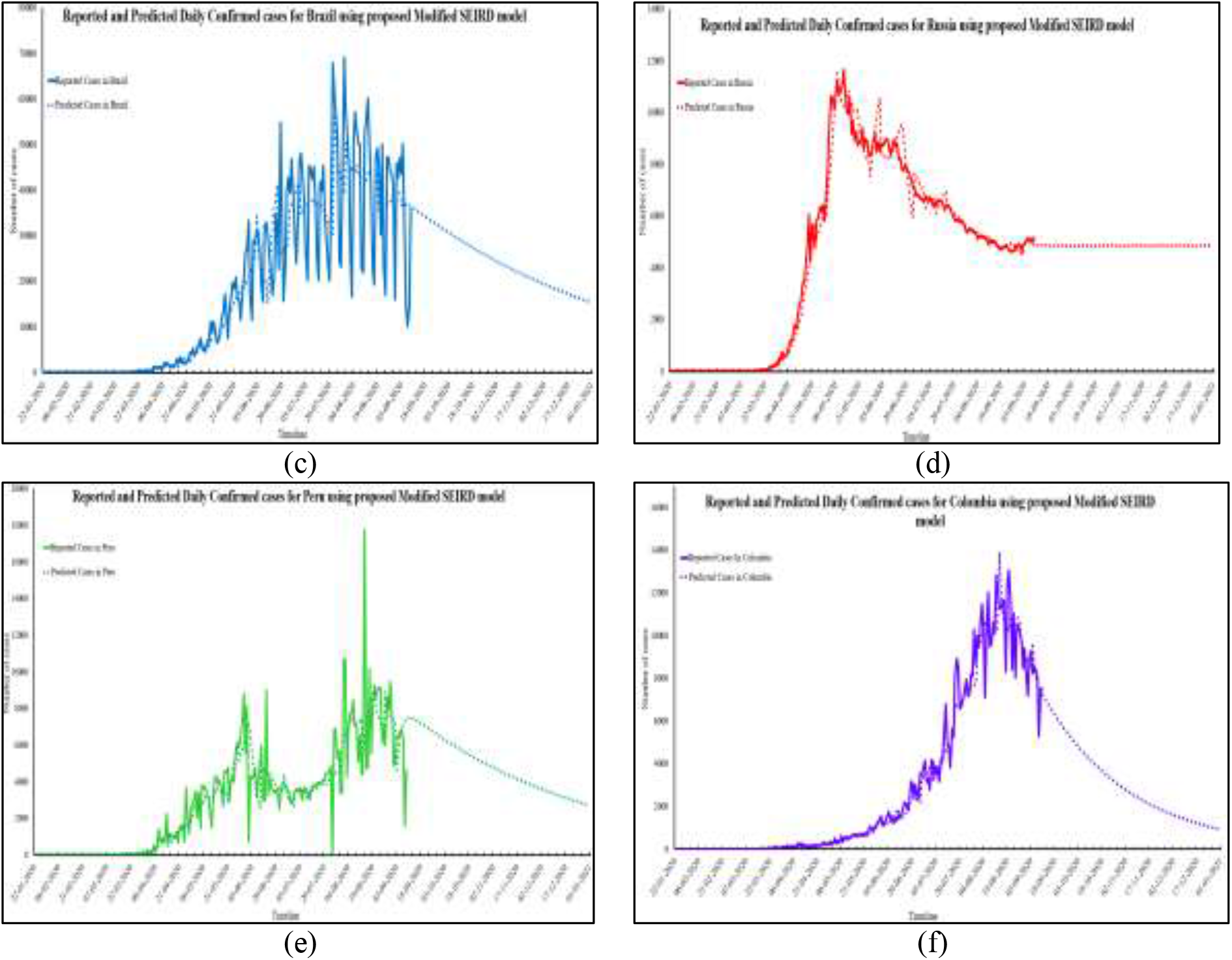
Reported and Predicted Daily Confirmed cases for six worst-affected countries

As shown in Figure 2 (a), USA will have 65,648 Daily Confirmed cases by the end of year 2020. In India, this number will rise to 5,60,016 at the same time. Though India currently ranks second in the worst-affected nations by COVID-19, but the number of Daily cases in this country are much higher than USA which is the worst-affected nation of the world. In case of Brazil, Peru and Columbia, the number of Daily Confirmed cases will be 15,434; 2,729 and 922 respectively on 31^st^ December 2020. The predicted values of Daily Confirmed cases fit closely with the reported data as shown in Figure 2 (a-f). This is also verified using the MAPE values calculated between the reported data and the predicted data. The MAPE values computed for Daily Confirmed cases of USA, India, Brazil, Russia, Peru and Colombia are 0.19, 0.22, 0.30, 0.13, 0.26 and 0.17 respectively. Figure 3 (a-f) presents the reported and predicted values for Daily Recovered cases of USA, India, Brazil, Russia, Peru and Colombia respectively.

**Figure 3(a-f):**
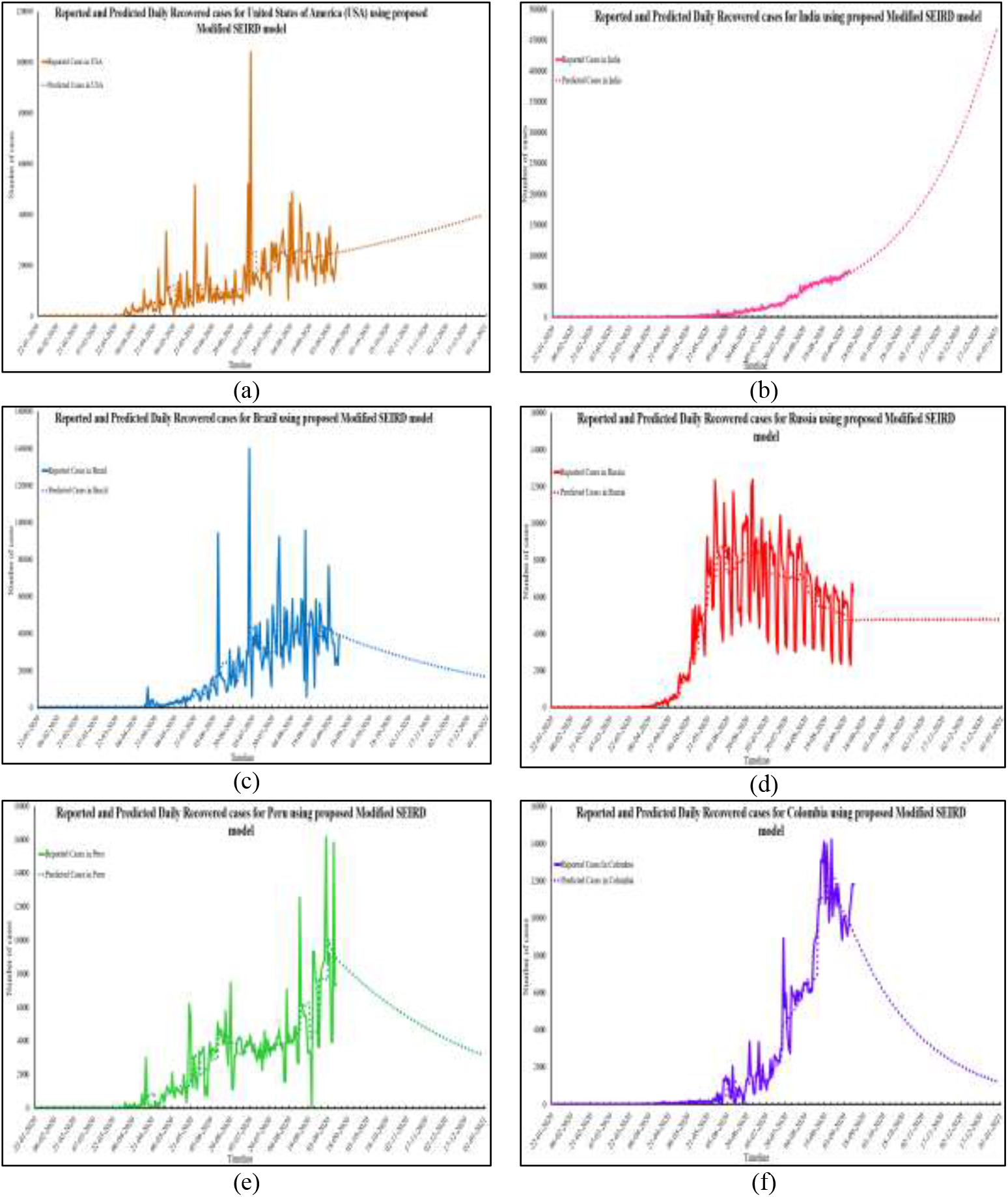
Reported and Predicted Daily Recovered cases for six worst-affected countries

The Modified SEIRD model predicts that by the end of year 2020, USA will have 40,014 Daily Recovered cases, whereas India, Brazil, Russia, Peru and Colombia will have 4,66,363; 16,679; 4,796; 3,152 and 1,179 Daily Recovered cases repectively. Among all the six countries in consideration, India has the highest number of Daily Recovered cases. The MAPE values obtained for Daily Recovered cases in case of USA, India, Brazil, Russia, Peru and Colombia are as follows: 1.22, 0.28, 0.44, 0.30, 0.81 and 2.71. For four out of six worst-affected nations, the MAPE values lie below 1, which indicates that the predictions are in close correspondence with the reported data. Figure 4 (a-f) shows the Daily Deceased cases for the six countries that have been worst affected by COVID-19.

**Figure 4(a-f):**
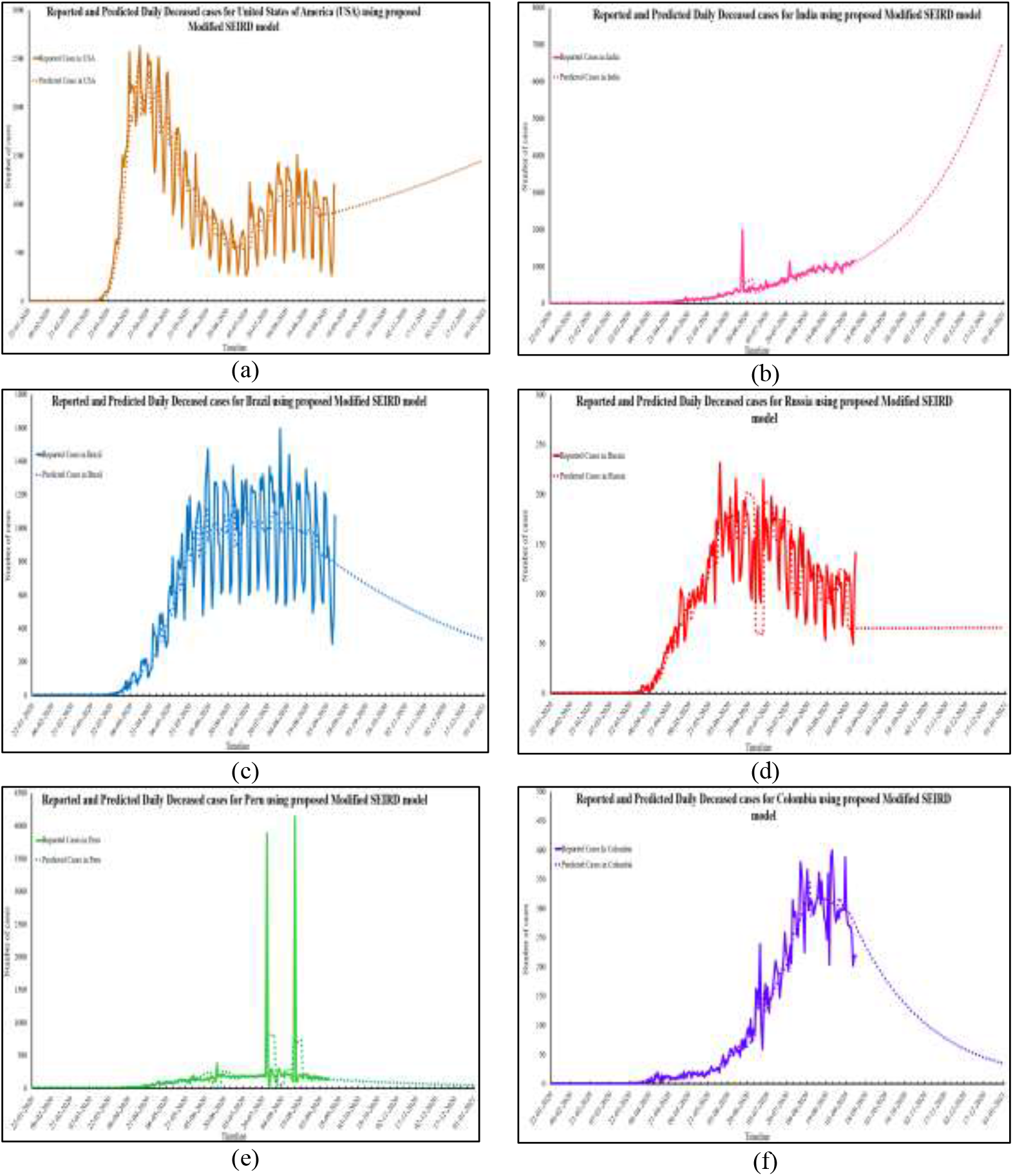
Reported and Predicted Daily Deceased cases for six worst-affected countries

The severity of any pandemic can be judged by the number of deaths caused by it. The COVID-19 pandemic has shown different CFR values in different countries. The predictions obtained from the Modified SEIRD model depict that USA will witness 1,456 deaths daily by the end of year 2020. Though the CFR value for India is very low, but the number of Daily Deceased cases in India will be 7,077 on 31^st^ December 2020. This can be attributed to the high rate of infection spread in this country. In contrast, Brazil, Russia, Peru and Colombia will have comparatively few Daily deaths caused by the infection. MAPE values calculated for Daily Deceased cases were found to be below 0.4 in all the cases. Figure 5 (a-f) show the Total Confirmed cases for worst affected six countries.

**Figure 5(a-f):**
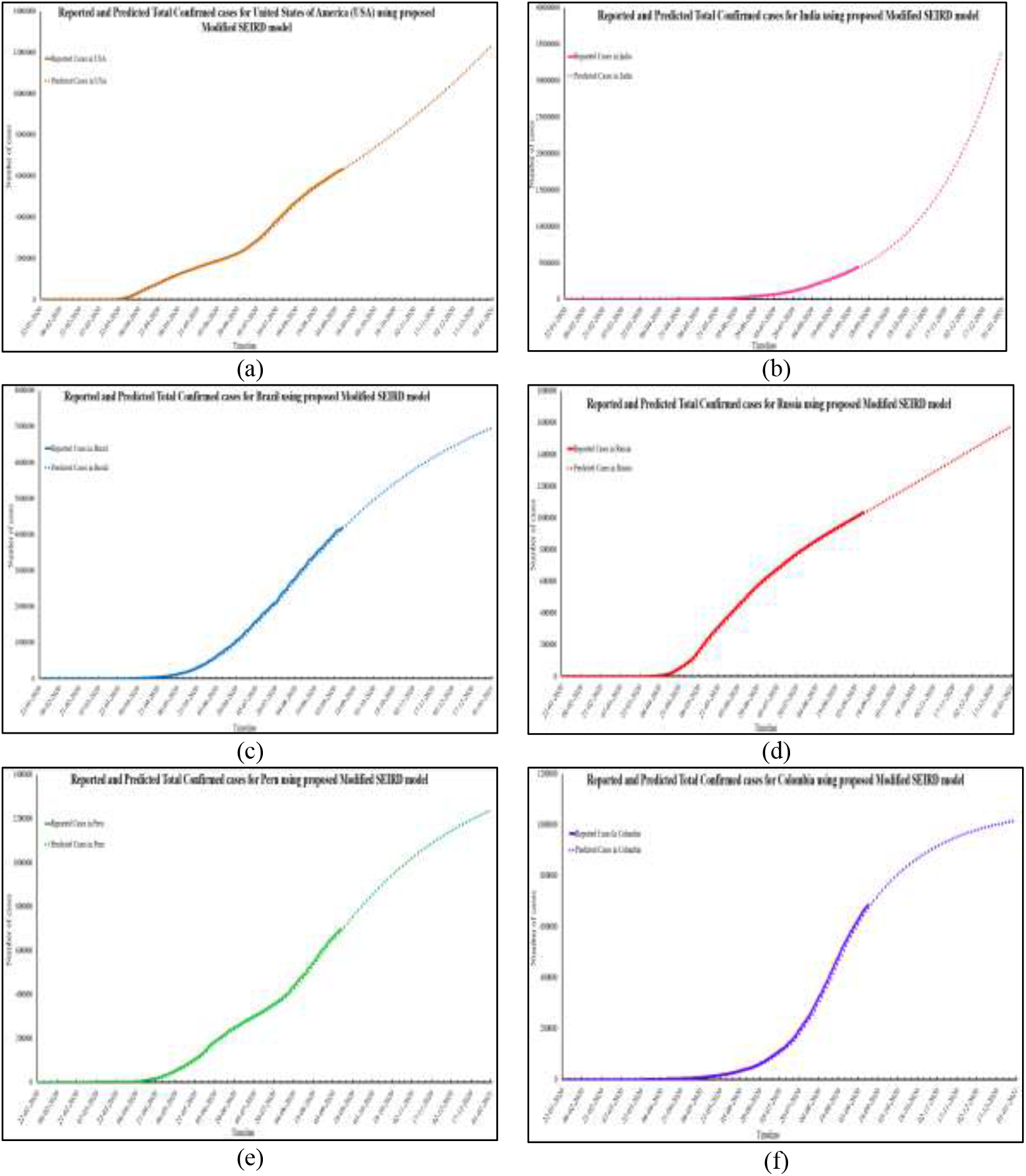
Reported and Predicted Total Confirmed cases for six worst-affected countries

The Modified SEIRD model predicts 1,23,37,873 Total Confirmed cases in USA as on 31^st^ December 2020. For India and Brazil, this number is expected to be 3,39,94,949 and 69,47,629 respectively. On the other hand, Russia will have 15,76,715 Total Confirmed cases while Peru will have 12,37,160 cases around the same time. In Colombia, the estimated number of Total Confirmed cases are 10,17,620. From these values, it can be concluded that by the end of 2020, India will have the highest number of Total Confirmed cases and will replace USA as the worst affected nation by COVID-19 pandemic. The MAPE values corresponding to Total Confirmed data, for all the six countries in consideration are 0.12, 0.16, 0.09, 0.24, 0.09 and 0.09 respectively. Next, Figure 6 (a-f) present the graphs of Total Recovered Cases for USA, India, Brazil, Russia, Peru and Colombia.

**Figure 6(a-f):**
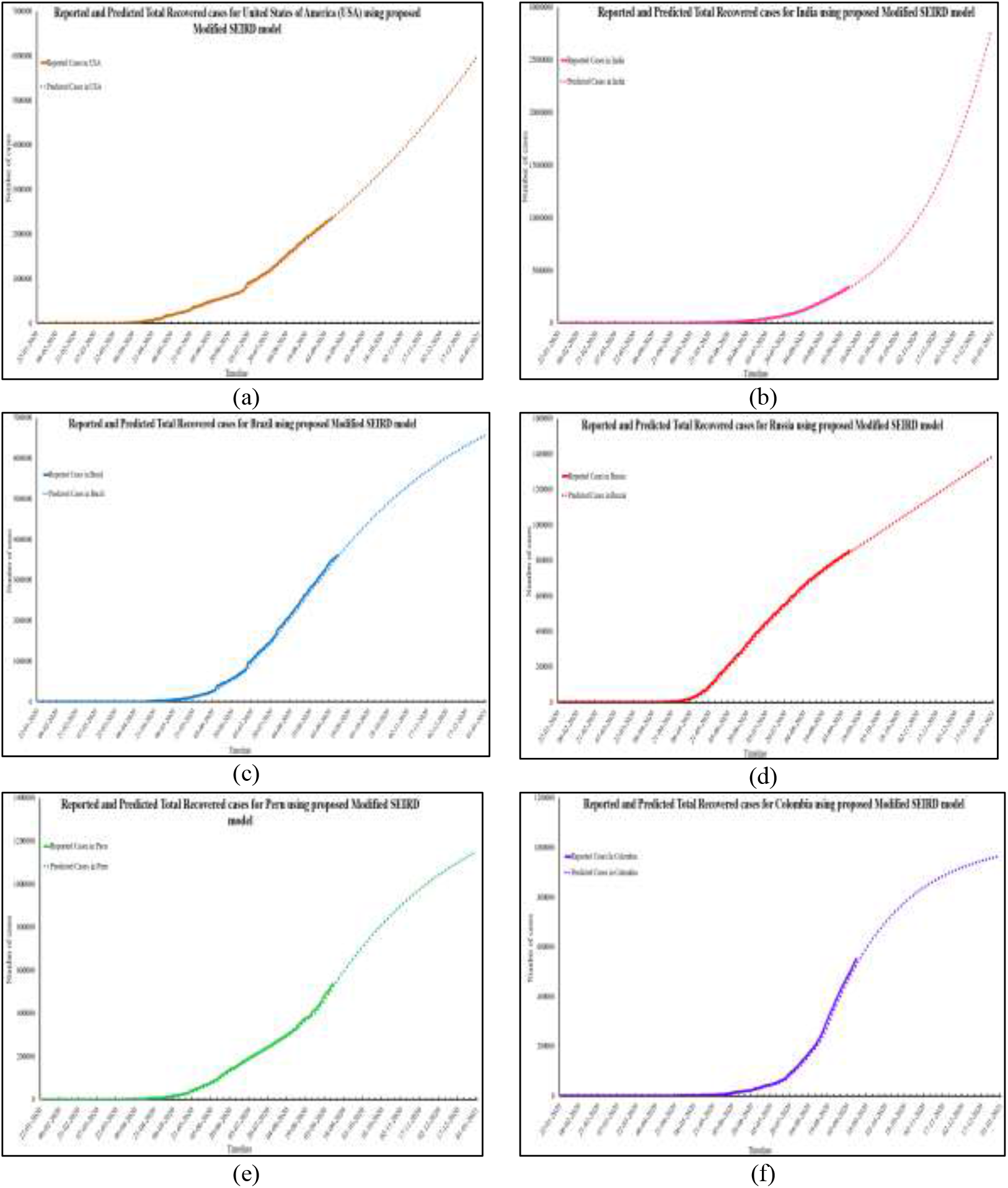
Reported and Predicted Total Recovered cases for six worst-affected countries

The Total Recovered cases for USA, India, Brazil, Russia, Peru and Colombia at the year end, will be 60,00,719; 2,78,23,877; 65,56,224; 13,84,586; 11,48,564 and 9,67,426. The MAPE values for Total Recovered cases in USA, India, Brazil, Russia, Peru and Colombia are 0.18, 0.14, 0.62, 0.15, 0.23 and 0.10 respectively. The low MAPE values indicate that the prediction made by the Modified SEIRD model closely correspond with the reported data with low error values for all the six nations in consideration. Figure 7 (a-f) represents the graphs for six worst-affected countries with Total Deceased time-series.

**Figure 7(a-f):**
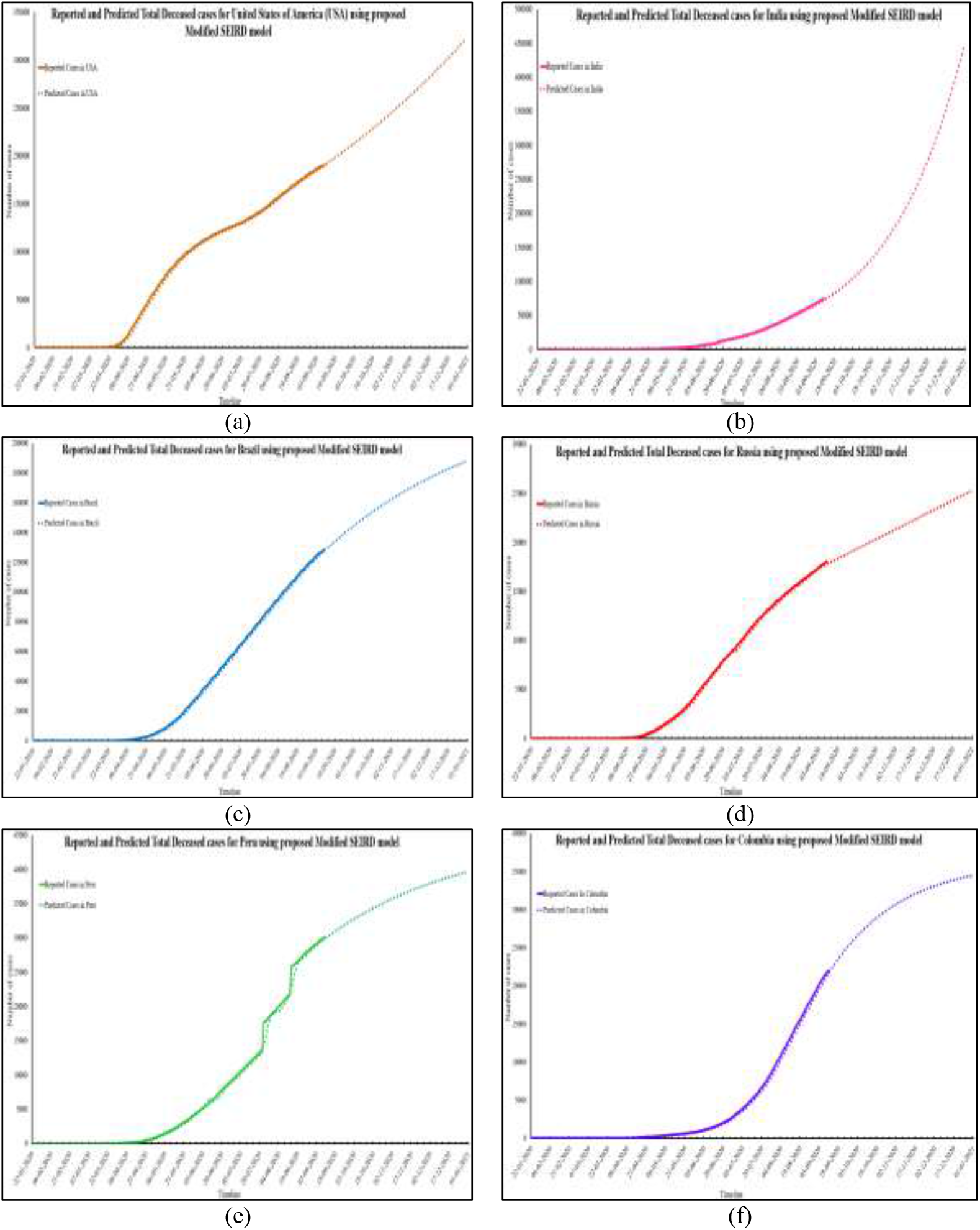
Reported and Predicted Total Deceased cases for six worst-affected countries

According to the Modified SEIRD model, USA and India will face 3,22,451 and 4,44,568 Total Deceased cases. Brazil, Russia, Peru and Colombia will have 1,87,855; 25,246; 39,650 and 34,522 Total Deceased cases at end of year 2020. The MAPE values corresponding to the same are: 0.09, 0.09, 0.08, 0.07, 0.07 and 0.07. Since the MAPE values for all the six worst-affected nations lie below 0.1, it can be safely concluded that the predictions made by the Modified SEIRD model are significantly similar to the reported data for Total Deceased cases. Next, Figure 8 (a-f) presents the Active Infected cases for the six worst-affected nations.

**Figure 8(a-f):**
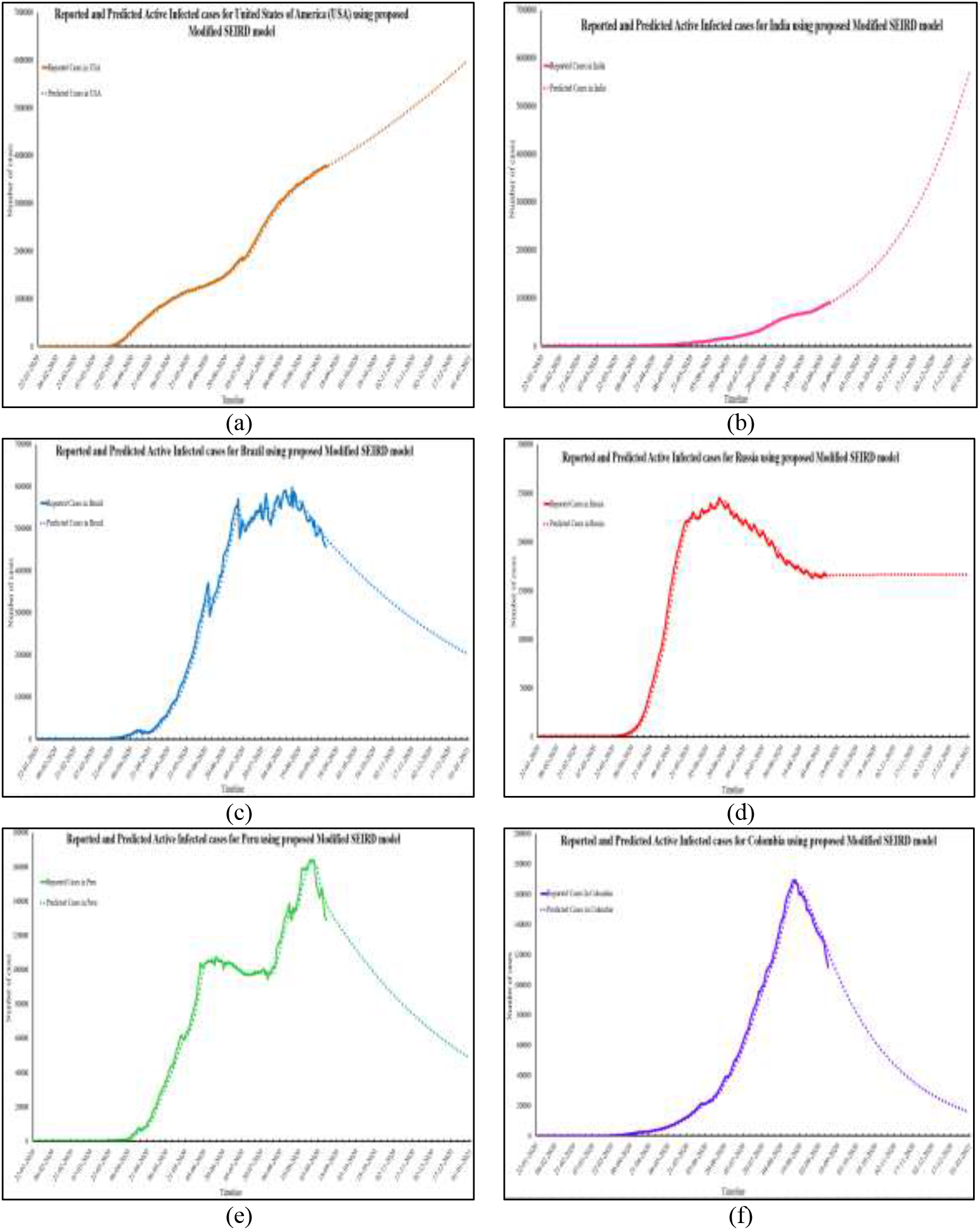
Reported and Predicted Active Infected cases for six worst-affected countries

The Modified SEIRD model predicts that by the end of year 2020, USA will have 60,14,704 Active Infected cases, whereas India, Brazil, Russia, Peru and Colombia will have 57,26,504; 2,03,550; 1,66,882; 48,946 and 15,672 Active Infected cases repectively. The MAPE values of Active Infected cases for USA, India, Brazil, Russia, Peru and Colombia are as follows: 0.11, 0.11, 0.12, 0.12, 0.08 and 0.09. For all the six worst-affected nations, the MAPE values lie below 0.2, which indicates that the predictions are in close correspondence with the reported data.

Further, statistical hypothesis testing using Student t-test was used for accepting the short-term predictions performed by the Modified SEIRD model. The reported and predicted values of Total Confirmed cases, Total Recovered cases, Total Deceased cases and Active Infected cases for USA, India, Brazil, Russia, Peru and Columbia have been used for performing t-test. The t-test was not performed for Daily cases due to fluctuations in daily data as the cases may not be reported timely. The t-test is used to perform a hypothesis test of the null hypothesis that the actual and predicted data are similar for short-term predictions. In this paper, paired t-test using two samples for means assuming unequal variance has been used. The null hypothesis and the alternate hypothesis are defined as follows:

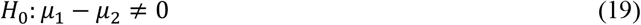

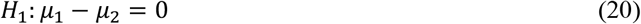

where, *μ*_1_ is the mean of the reported data and *μ*_2_ is the mean our model in consideration.

T-test is used to estimate whether the observed data belongs to the null hypothesis or not. If the observed test statistic differs significantly from the hypothesized values, then null hypothesis is rejected and we claim the desired performance of the proposed algorithm by accepting the alternate hypothesis. For the experimentation, significance level as 0.01 was selected. In statistical hypothesis testing using t-test, probability p is calculated. If the value of p is less than 0.01, then the null hypothesis is rejected and alternate hypothesis is accepted with 99% confidence. Otherwise, null hypothesis is rejected and alternate hypothesis is accepted.

For Total Confirmed cases, the p-values obtained for six worst affected countries of world including USA, India, Brazil, Russia, Peru and Colombia were less than 0.01. In case of Total Recovered predictions, all the p-values were below 0.01 significance level. Similarly, the p-value of Total Deceased and Active Infected cases are also less than 0.01. Therefore, in all these cases, the null hypothesis is rejected and the alternate hypothesis is accepted with 99% confidence. The above-mentioned results show that the predictions of the Modified SEIRD model closely correspond with the reported data. Hence, the Modified SEIRD model is further used to perform long-term predictions of COVID-19 in six countries in consideration. The results for the same are presented in the next section.

### 5.2. Results for Long Term Prediction

The long-term predictions were performed till the end of year 2023 for the six worst-affected nations being considered in this paper. The predictions for different time-series data obtained from the Modified SEIRD model have been discussed below. The Effective Reproduction Number, Re, has also been calculated and analysed. For calculating Re, the starting date has been selected as 1^st^ April 2020 as by this time, the number of reported cases in all the six countries in consideration surpassed 1000 infected cases (Linka, Peirlinck, & Kuhl, 2020). This has been shown in Figure 9 (a-c) - Figure 14 (a-c).

**Figure 9(a):**
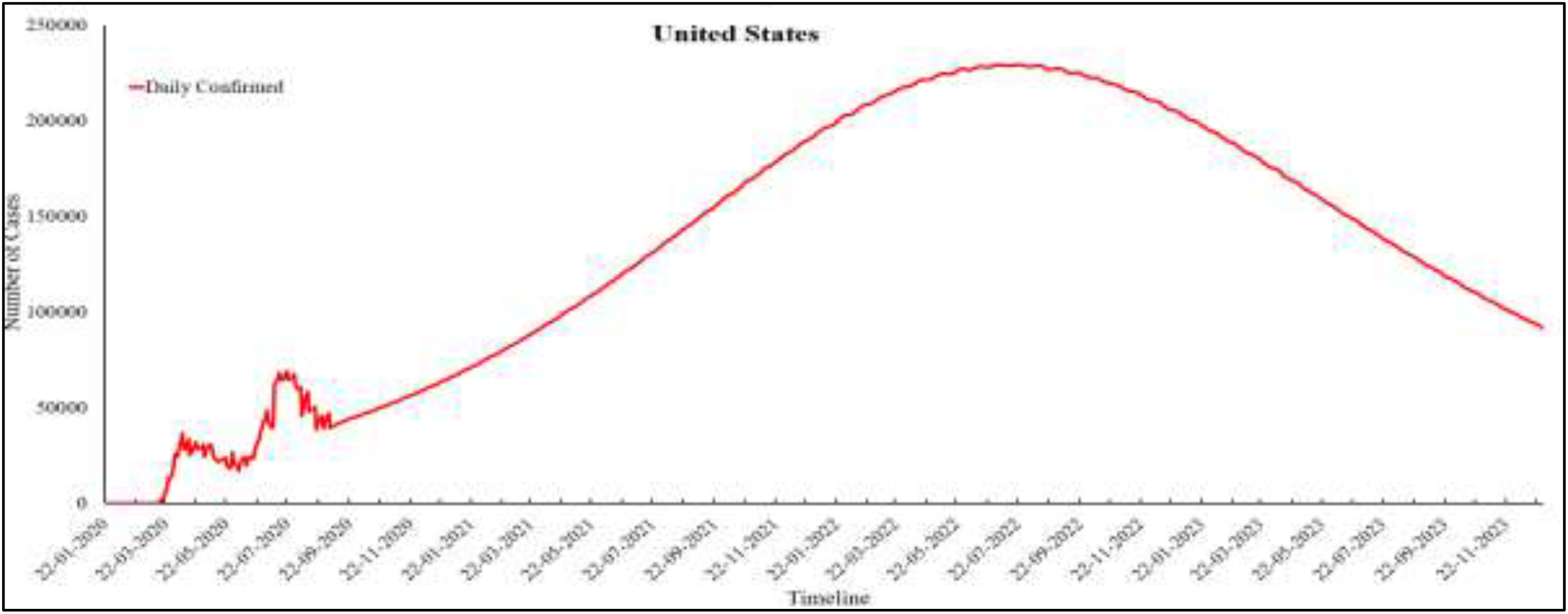
Predicted Daily Confirmed cases for USA

**Figure 9(b):**
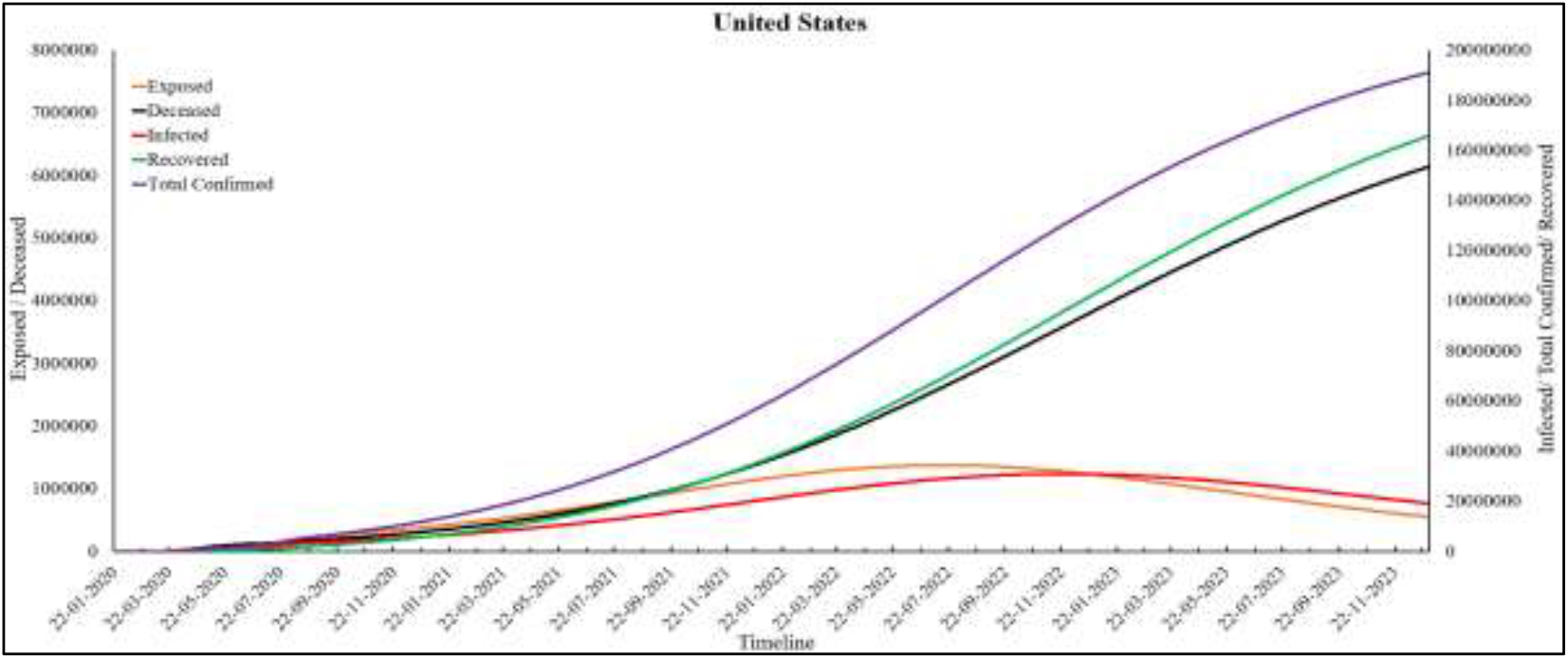
Predictions for USA

**Figure 9(c):**
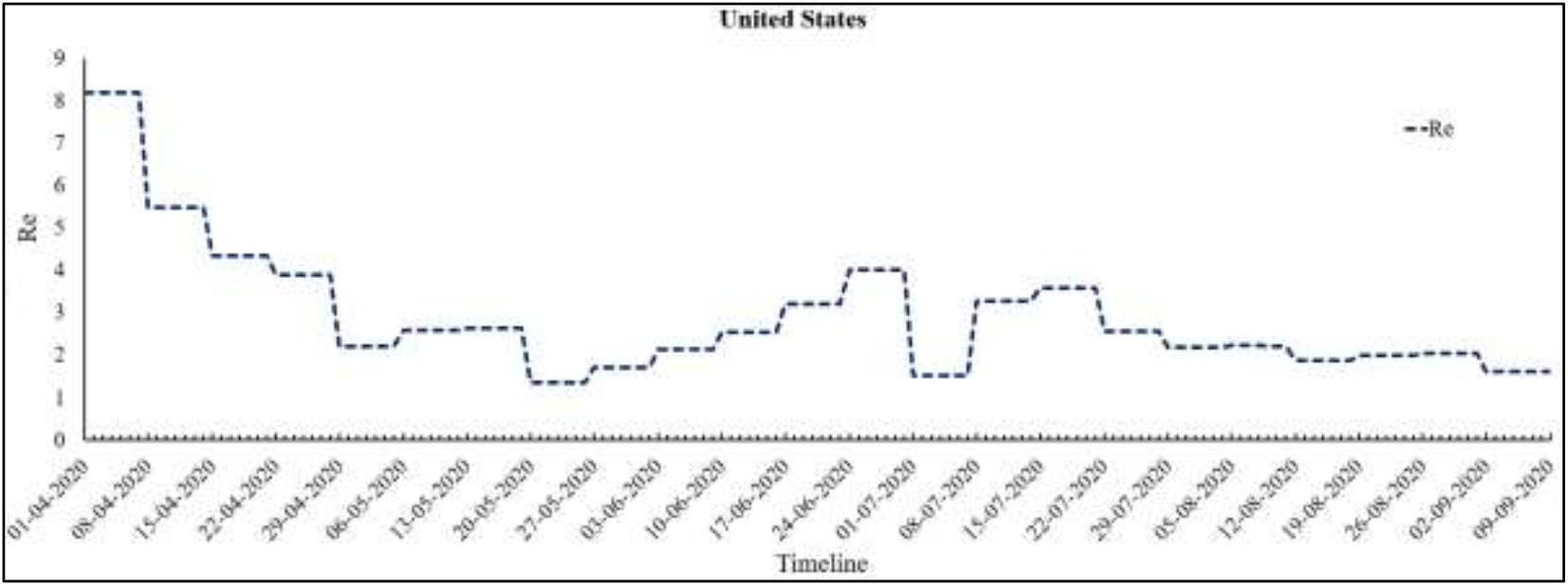
Re values for USA

**Figure 10(a):**
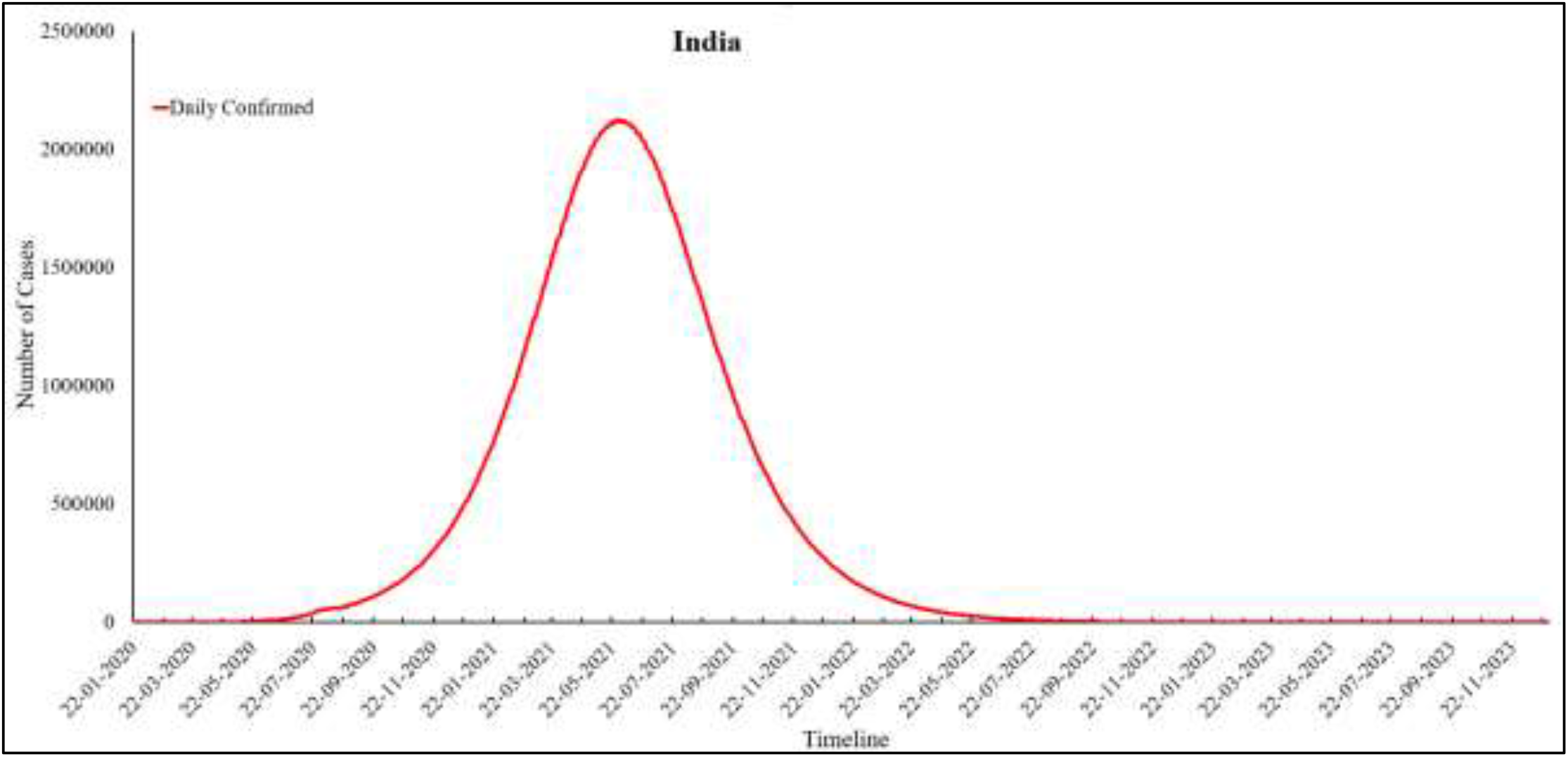
Predicted Daily Confirmed cases for India

**Figure 10(b):**
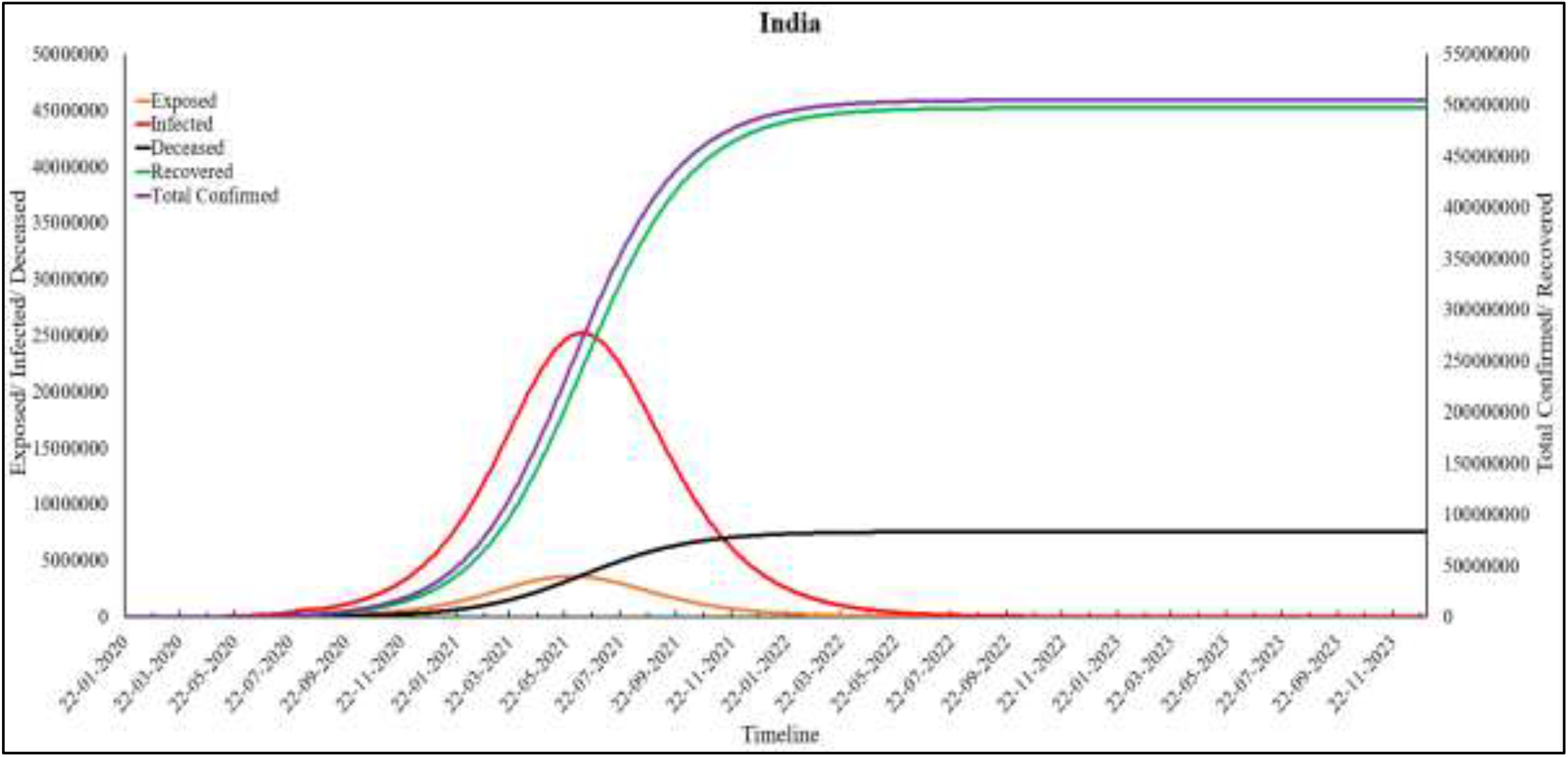
Predictions for India

**Figure 10(c):**
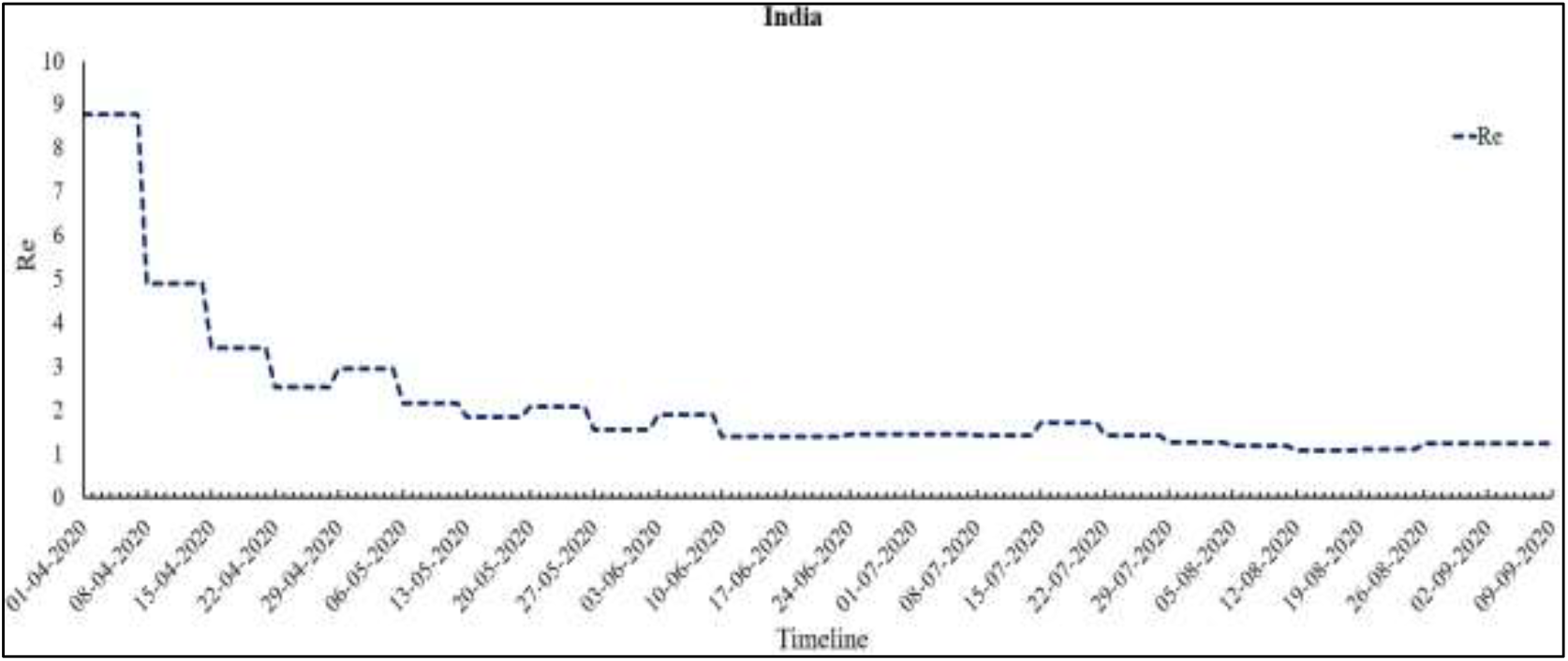
Re values for India

**Figure 11(a):**
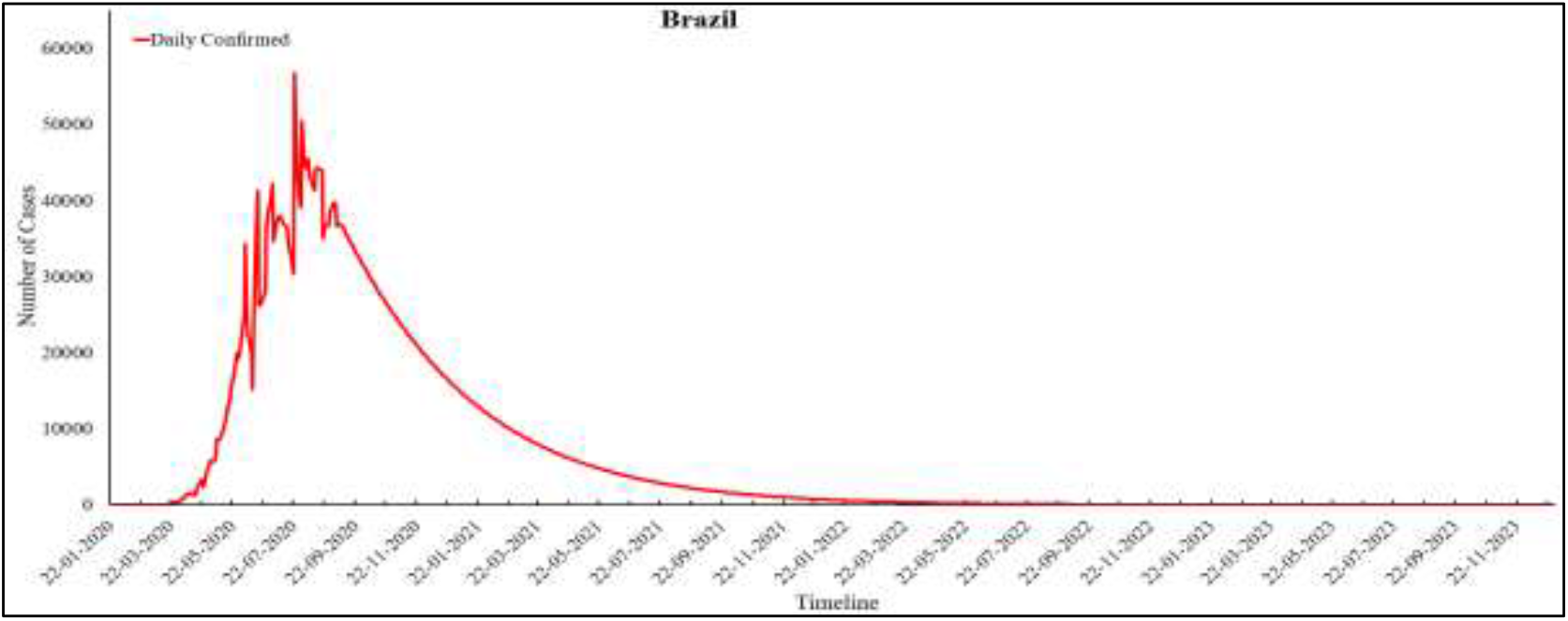
Predicted Daily Confirmed cases for Brazil

**Figure 11(b):**
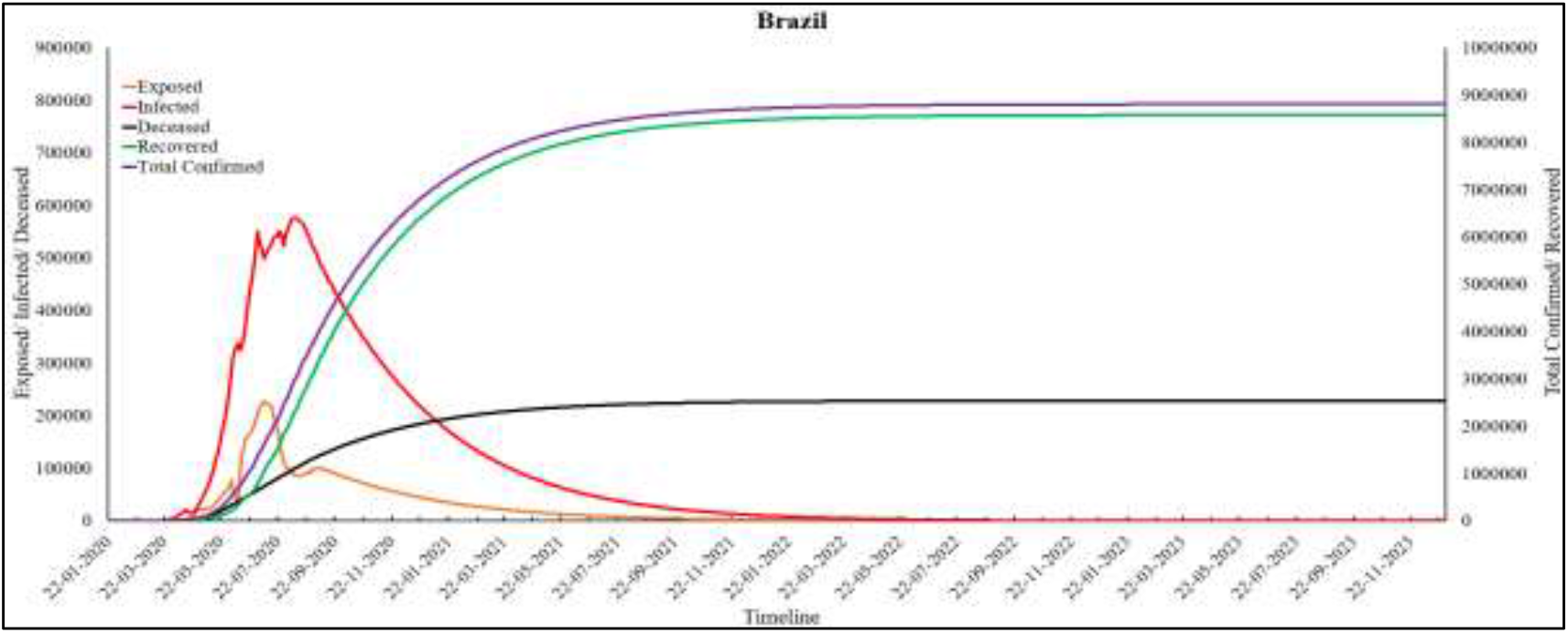
Predictions for Brazil

**Figure 11(c):**
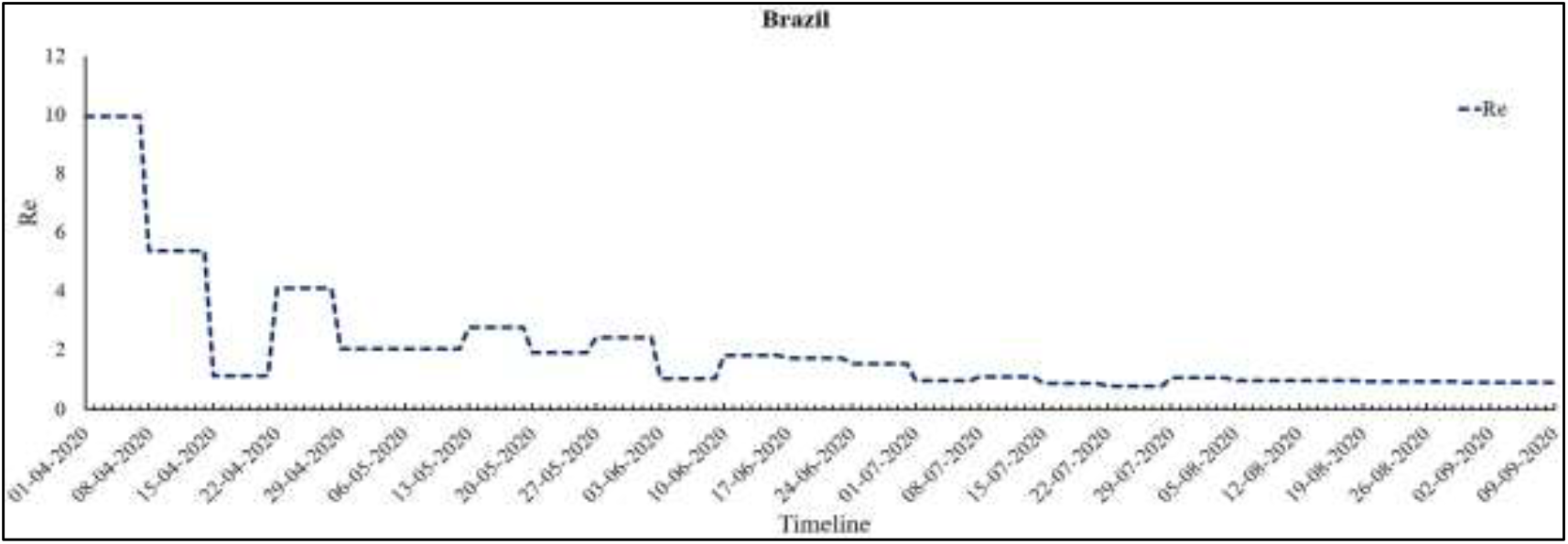
Re values for Brazil

**Figure 12(a):**
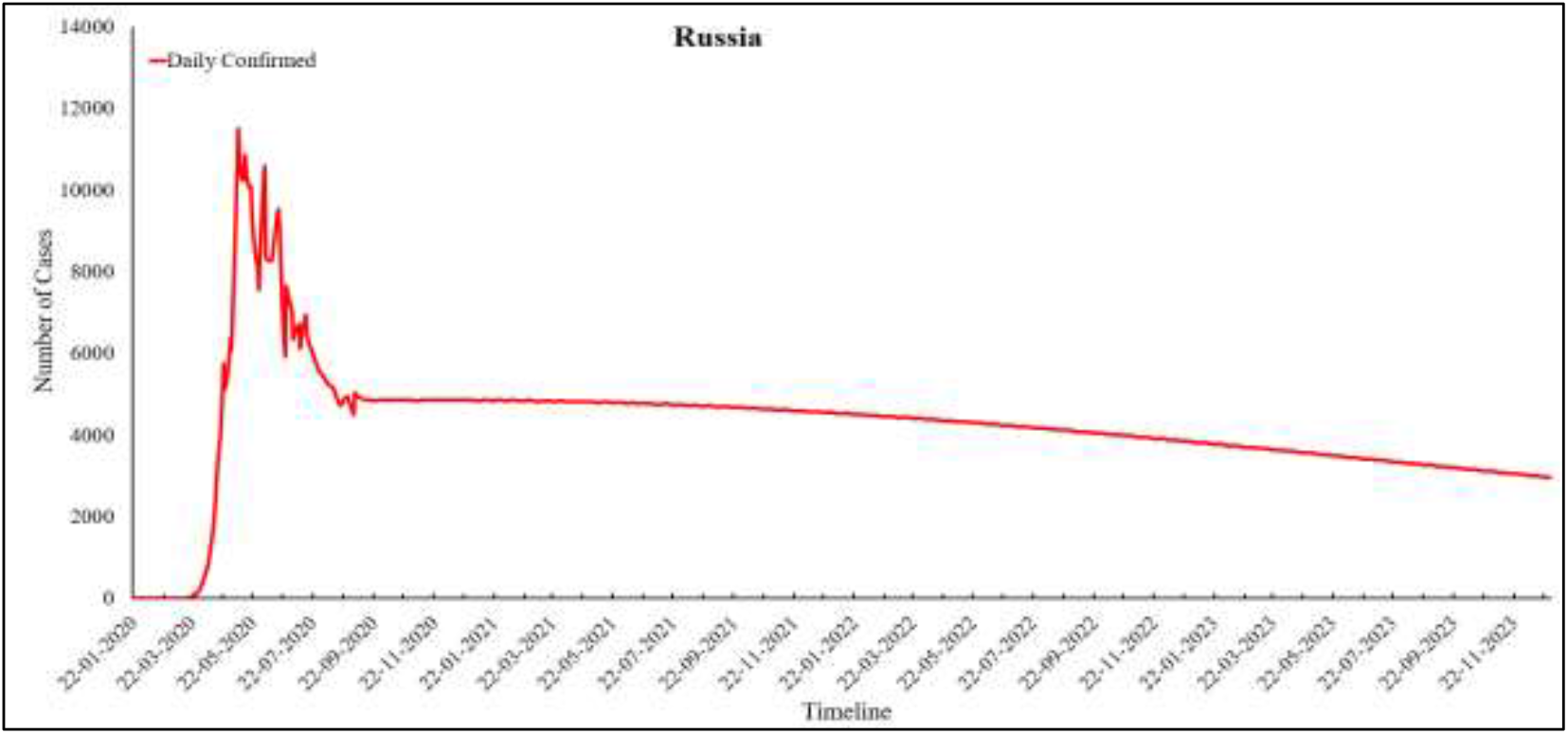
Predicted Daily Confirmed cases for Russia

**Figure 12(b):**
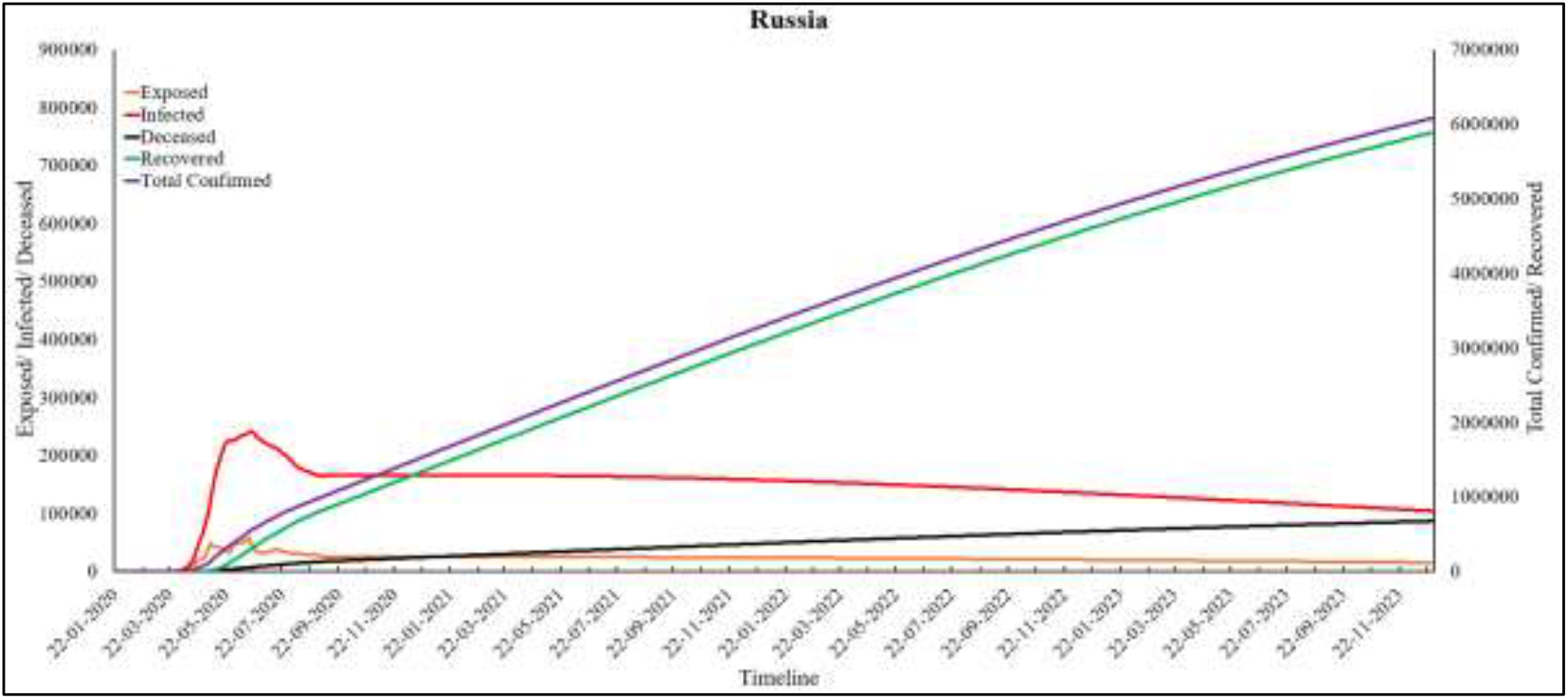
Predictions for Russia

**Figure 12(c):**
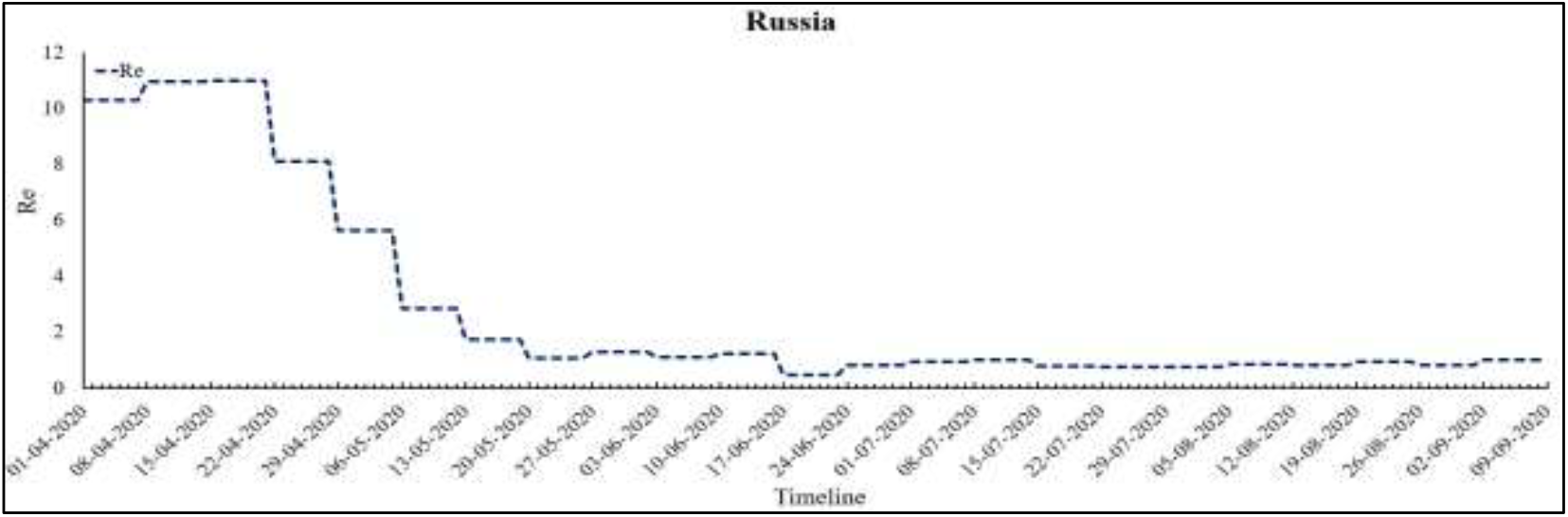
Re values for Russia

**Figure 13(a):**
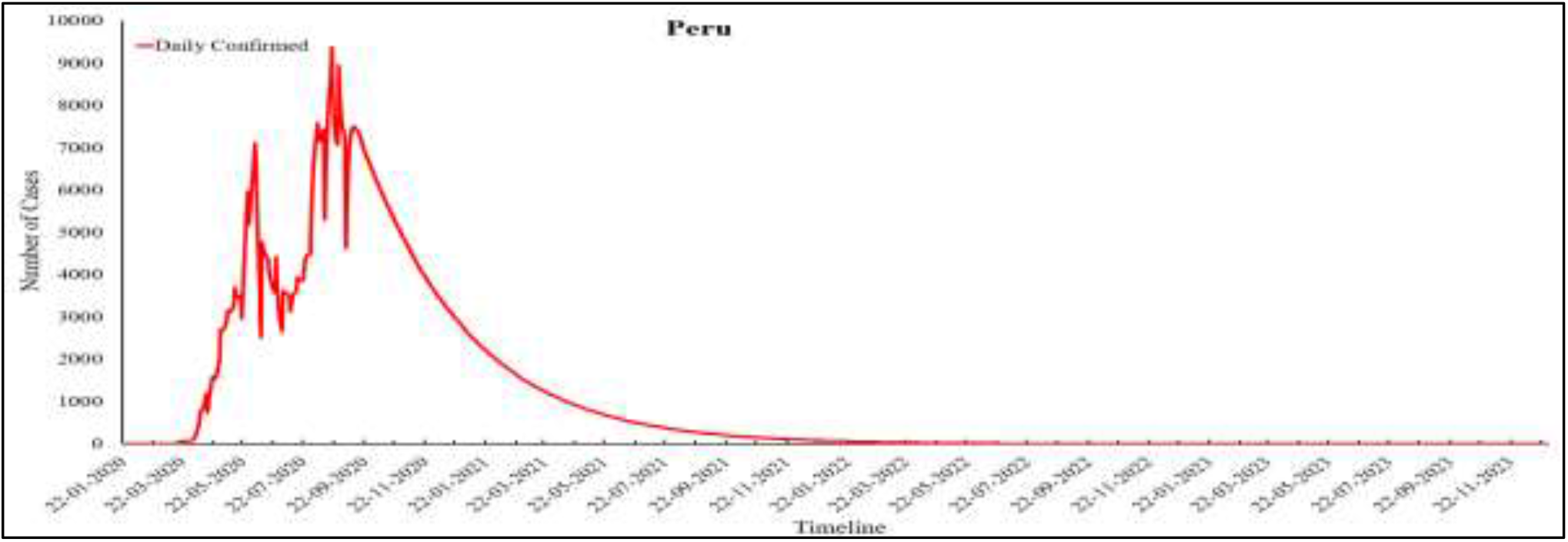
Predicted Daily Confirmed cases for Peru

**Figure 13(b):**
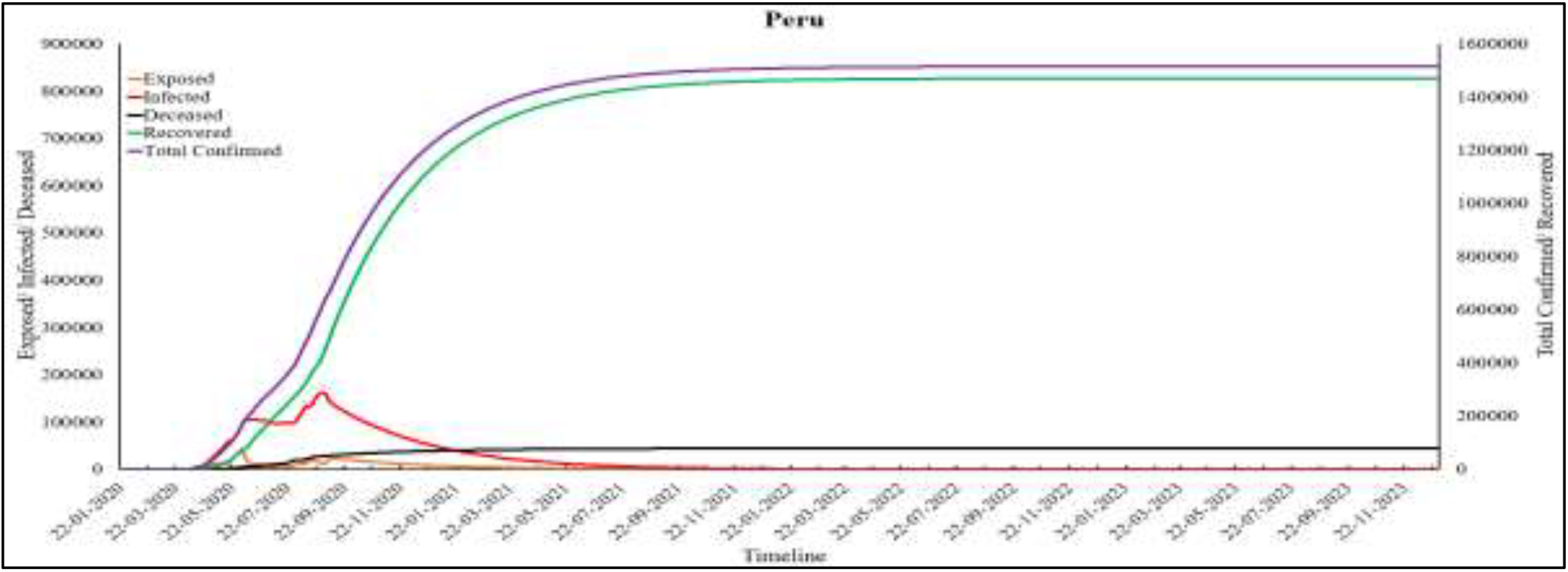
Predictions for Peru

**Figure 13(c):**
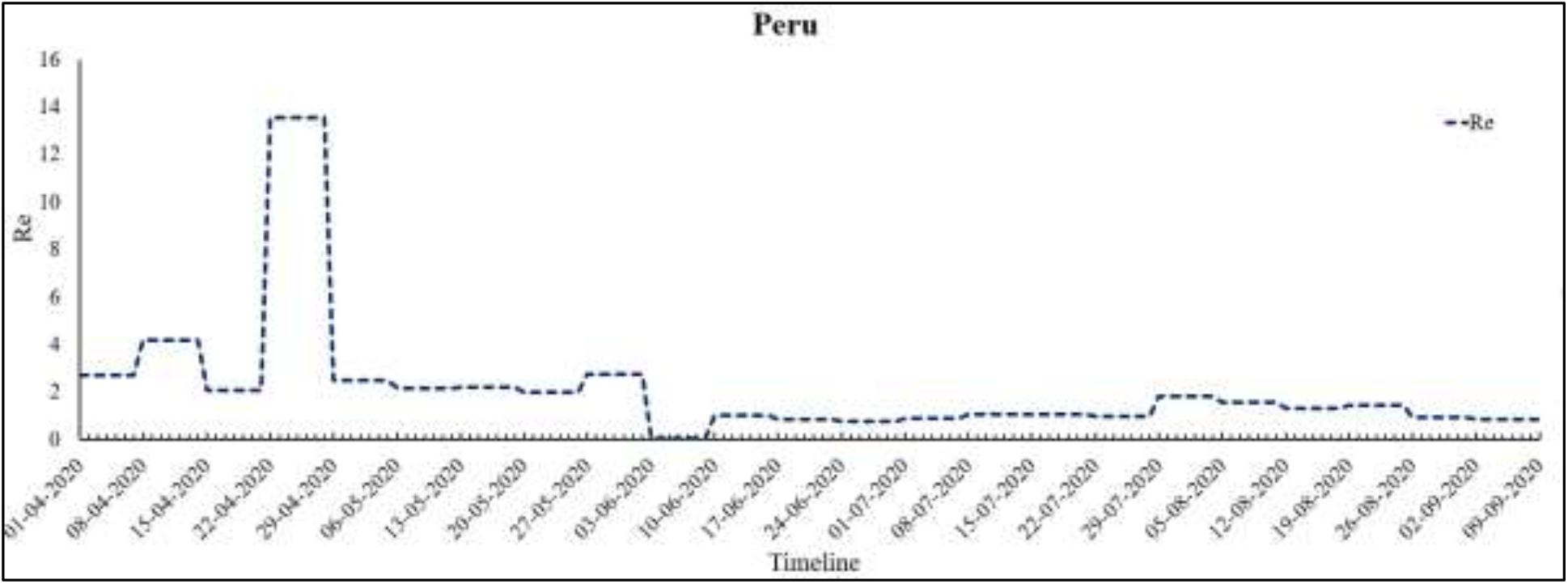
Re values for Peru

**Figure 14(a):**
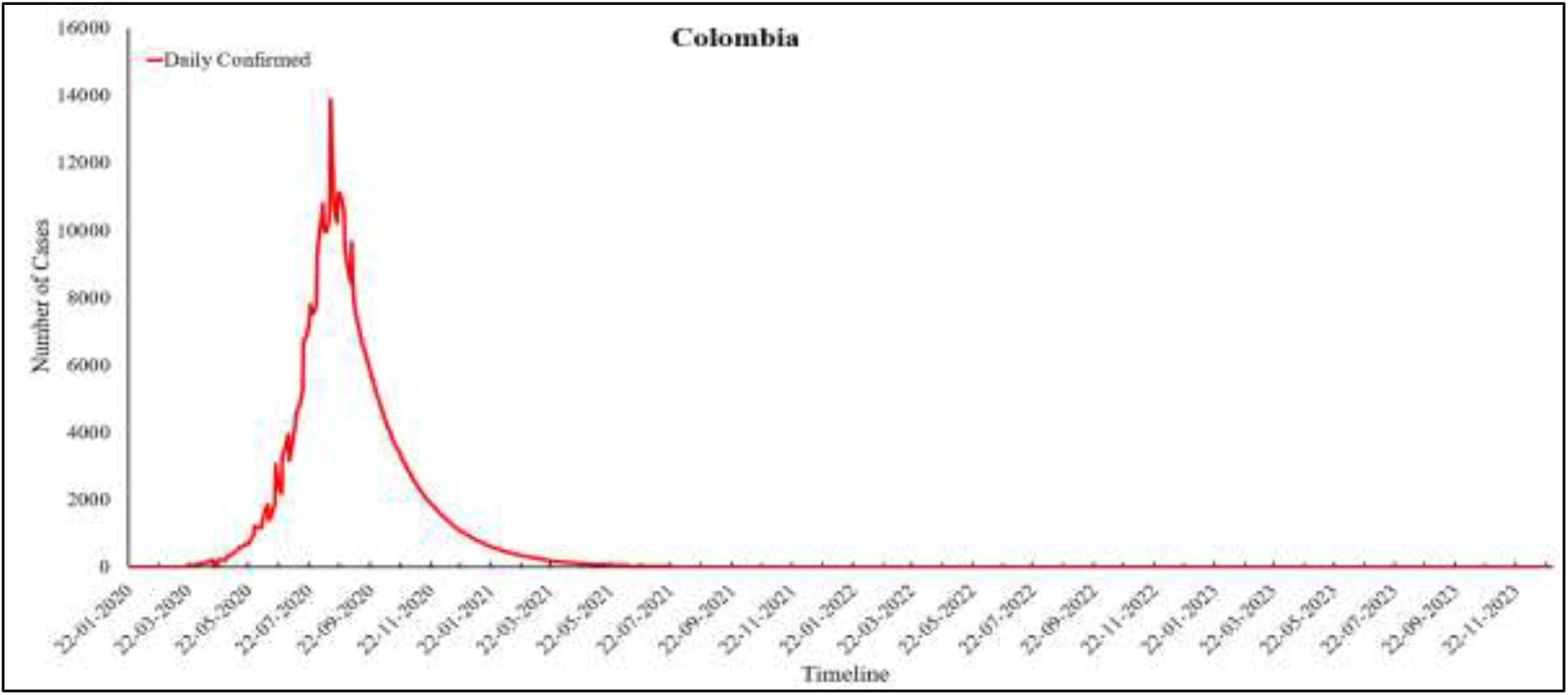
Predicted Daily Confirmed cases for Colombia

**Figure 14(b):**
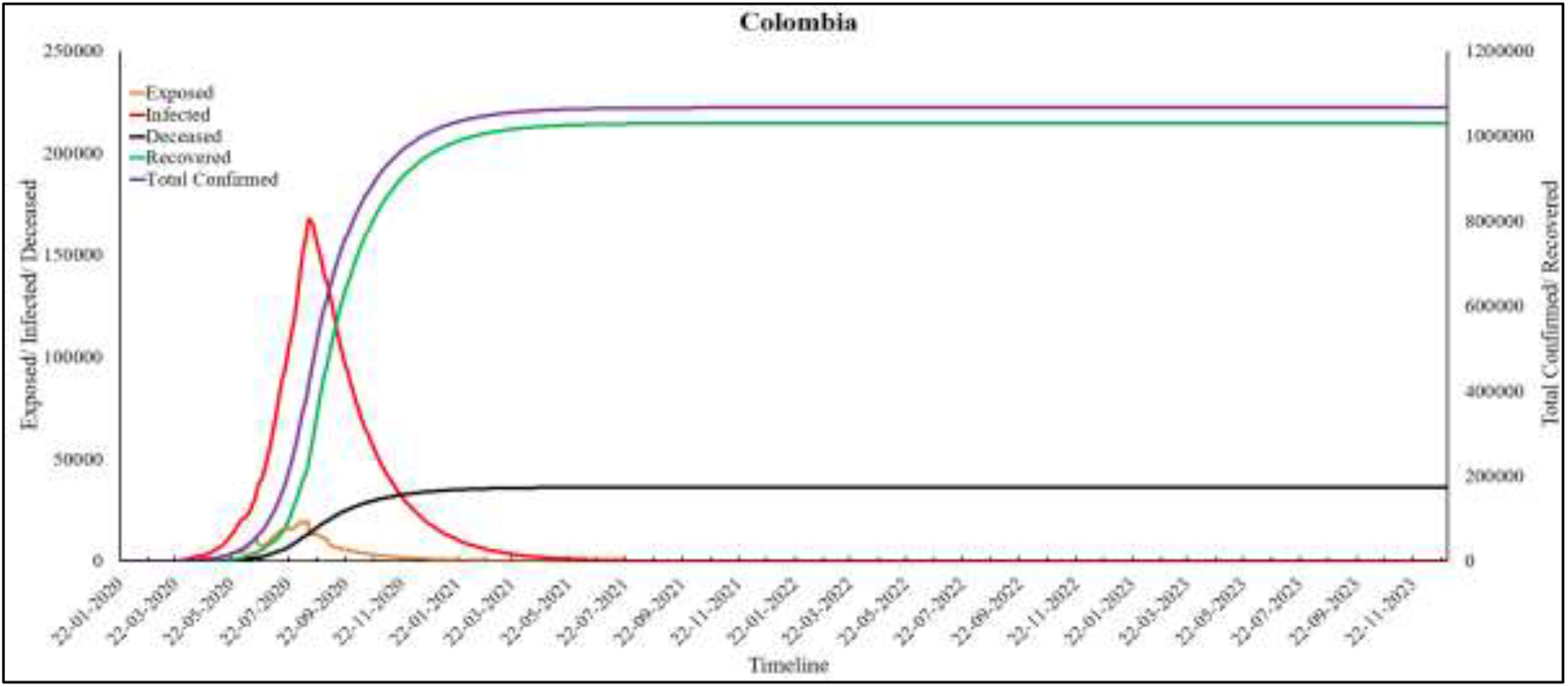
Predictions for Colombia

**Figure 14(c):**
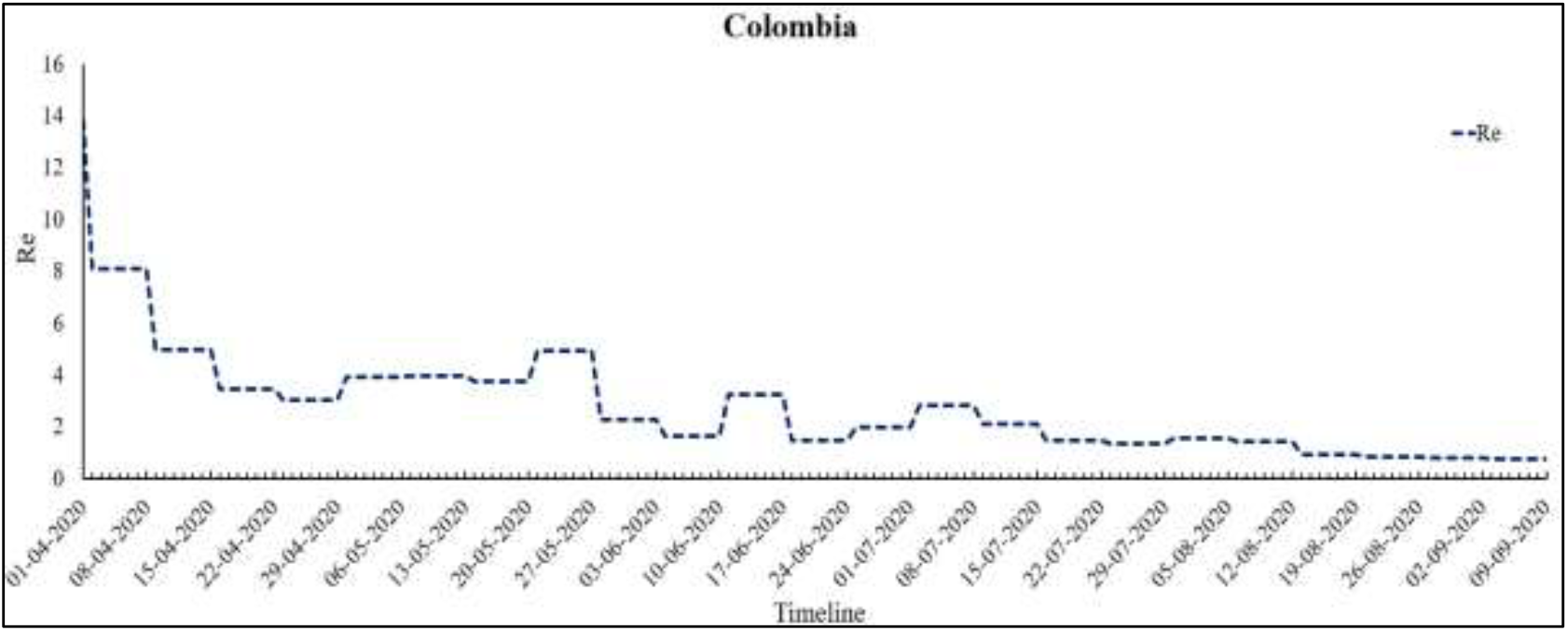
Re values for Colombia

#### United States of America (USA)

In USA, first case of COVID-19 was encountered in January and as on 9^th^ September 2020, this number increased to 6,361,265 (Worldometer, 2020). In the initial phase, the US government downplayed the severity of virus and did not take necessary precautions to control the spread. Therefore, the value of β which represents the transmission rate per capita, reached 0.37 at the beginning of March and by mid-March, it reached 0.49. At the same time, the value of ε was 0.99, which reflects that the infectious exposed portion of population is very high in USA. Therefore, a national emergency was declared in USA in March itself (Liptak, 2020). The authorities started purchasing medical equipments in bulk, stay at home orders were released and schools were closed. In April, the guidelines were updated and a new division named as Coronavirus Commission for Safety and Quality in Nursing Homes was setup (Whitehouse.gov, 2020). In the same month, USA achieved the highest CFR by crossing Italy, as the value of mu increased from 0.0053 to 0.0059. In June, a second wave of infection was seen when the relaxations were given in some states and again the value of β increased from 0.01 to 0.03. On 1^st^ April 2020, the Re value of USA was estimated to be 8.18 which has reduced to 1.61 as on 9^th^ September 2020. According to the Modified SEIRD model, USA will see its peak in July 2022. The Daily Confirmed cases at the time of peak will be 2,29,457 and Total Confirmed cases will be 10,25,02,451. The Total Deceased cases will be 26,74,542 whereas the Total Recovered cases will be 7,06,24,200. At the same time, the number of Active Infected cases in USA will be 60,14,704. These values can be observed from Figure 9 (a-c).

#### India

India is a diverse nation with a huge population and high population density. In the beginning of March, the value of β was found to be very high which was 0.77. The government of India responded promptly by locking the whole country in the early stages of the pandemic. This lockdown continued in four stages, starting from 25^th^ March 2020 to 31^st^ May 2020. After lockdown 1.0, there was a decrease in transmission rate (β) from 0.34 to 0.17. During lockdown 4.0 which happened in May, the value of ε decreased to 0.0001, therefore the lockdown was lifted by giving relaxations and it was named as Unlock 1.0. There was an increasing trend observed in the value of ε in Unlock 1.0 period. In India, the Recovery Rate is very high as compared to other countries and a constant increase is observed in the value of *γ*. Also, the low CFR contributes to decreasing mu value in India. On 1^st^ April 2020, the Re value of India was estimated to be 8.79 which has reduced to 1.24 as on 9^th^ September 2020. The peak of India is expected to be in May 2021 with 21,26,406 cases coming daily. The Total Confirmed cases at peak will be 24,82,39,581 in which 89% population will be recovered and 0.01% will lose their fight against the infection. At this time, the number of Active Infected cases in India will be 57,26,504. These values can be observed from Figure 10 (a-c).

#### Brazil

Brazil has the third-highest number of Total Confirmed cases. Brazil had its first case of COVID-19 at the end of February and by the end of March, the infection had spread to most of the states in Brazil. In the month of March, the value of β increased to 0.888 from 0.001. The government of Brazil restricted air travel to control the spread of the pandemic. But in mid-April, the value of ε increased to 0.999. The reason for the same may be attributed to the relaxations in the social distancing norms given by the newly appointed Health minister, Nelson Luiz Sperle Teiz (G1, 2020). At the end of April, it was observed that the CFR increased in Brazil, therefore, the value of µ also increased from 0.006 to 0.013. According to the predictions obtained from the Modified SEIRD model, Brazil has already witnessed peak number of infected cases on 23^rd^ July 2020. The number of Active Infected cases at the time of peak were 2,03,550. On 1^st^ April 2020, the Re value of Brazil was found to be 9.92 which has reduced to 0.89 as on 9^th^ September 2020. Re value below 1 denotes a decreasing trend of the pandemic in the country. The end of pandemic is expected by the end of year 2023. The long-term predictions for Brazil have been shown in Figure 11(a-c).

#### Russia

The largest country in the world in terms of area, is ranked fourth place with 10,37,526 Total Confirmed cases as on 9^th^ September 2020. In Russia, the first case was found in February after the evacuation of Russian passengers from a cruise named Diamond Princess. This cruse was considered to be second largest hotbed of COVID-19 outside China (TASS, 2020). The value of ε from March to May was consistently high, showing that the infection has spread to a large section of the population. Recently, the value of *γ* has shown a decreasing trend from 0.36 to 0.28, whereas, the value of mu on 9^th^ September 2020 was found to be 0.0004. A low value of mu indicates a decline in the number of fatalities caused by the infection, which is the prime aim of the government in case of any pandemic. On 1^st^ April 2020, the Re value of Russia was estimated to be 10.29 which has reduced to 1 as on 9^th^ September 2020. This shows that the pandemic is on the decline in this country. However, Russia witnessed its infection peak on 7^th^ May 2020 but recently, a second wave of infection has been observed and the same has been confirmed by an increase in the value of Re. The pandemic in Russia is expected to be for a large period. The end of pandemic is expected even after the year 2023. The long-term predictions obtained by the Modified SEIRD model for Russia have been shown in Figure 12 (a-c).

#### Peru

On 6^th^ March 2020, the first case of COVD-19 was detected in Peru (Aquin & Garrison, 2020). After this, the government of Peru had put up several restrictions including lockdown, quarantine, and use of face masks in public areas. But people did not abide by these regulations, therefore, the infection spread at a high rate which is also reflected by the high value of ε. At the same time, Peru had the highest CFR, as shown by the high value of mu. On 1^st^ April 2020, the Re value of Peru was estimated to be 2.64 which has reduced to 0.84 as on 9^th^ September 2020. The peak of Peru was witnessed on 20^th^ August 2020 and the number of Active Infected cases at this time will be 48,946. The end of pandemic in Peru will be towards end of 2023. The long-term predictions for Peru has been shown in Figure 13 (a-c).

#### Colombia

Colombia is the sixth nation that has been most-affected by COVID-19 pandemic. This country witnessed its peak on 13^th^ August 2020 with 13,907 Daily cases and 15,672 Active Infected cases. On 1^st^ April 2020, the Re value of Colombia was estimated to be 13.83 which has reduced to 0.77 as on 9^th^ September 2020. The value of ε was found to be consistently high. The reason for the same can be attributed to the relaxations given by the government with high number of exposed asymptomatic individuals. Colombia observes a mix of infection transmission rate in the value of beta. The country has a relatively high CFR which is also shown by the value of mu. The long-term predictions for Colombia has been shown in Figure 14 (a-c).

The values of parameters (β, ε, α, γ and µ) optimized by the Modified SEIRD model have been presented in Table 3-7. The optimized values for the β parameter corresponding to the six nations in consideration, have been shown in Figure 14. For any pandemic, the β parameter reflects the rate at which susceptible people get infected. High β values correspond to high transmission rate per capita, indicating a fast spread of the infection. As can be seen from the table, India shows a high β value as compared to all other countries. In such cases, contact tracing must be increased and measures like social-distancing and isolation must be implemented strictly. For Brazil, high β values were obtained by the model till June, but later, a decline in values was observed. This corresponds to the actual scenario of COVID-19 in Brazil, as the peak for the infection was achieved around June. On the other hand, Russia showed high β values in the beginning and few spikes later on, but a downward trend was observed in the second half of the year 2020. Lastly, Peru and Columbia both show comparatively slow transmission rates of the COVID-19 infection as compared to their counterparts.

**Table 3:**
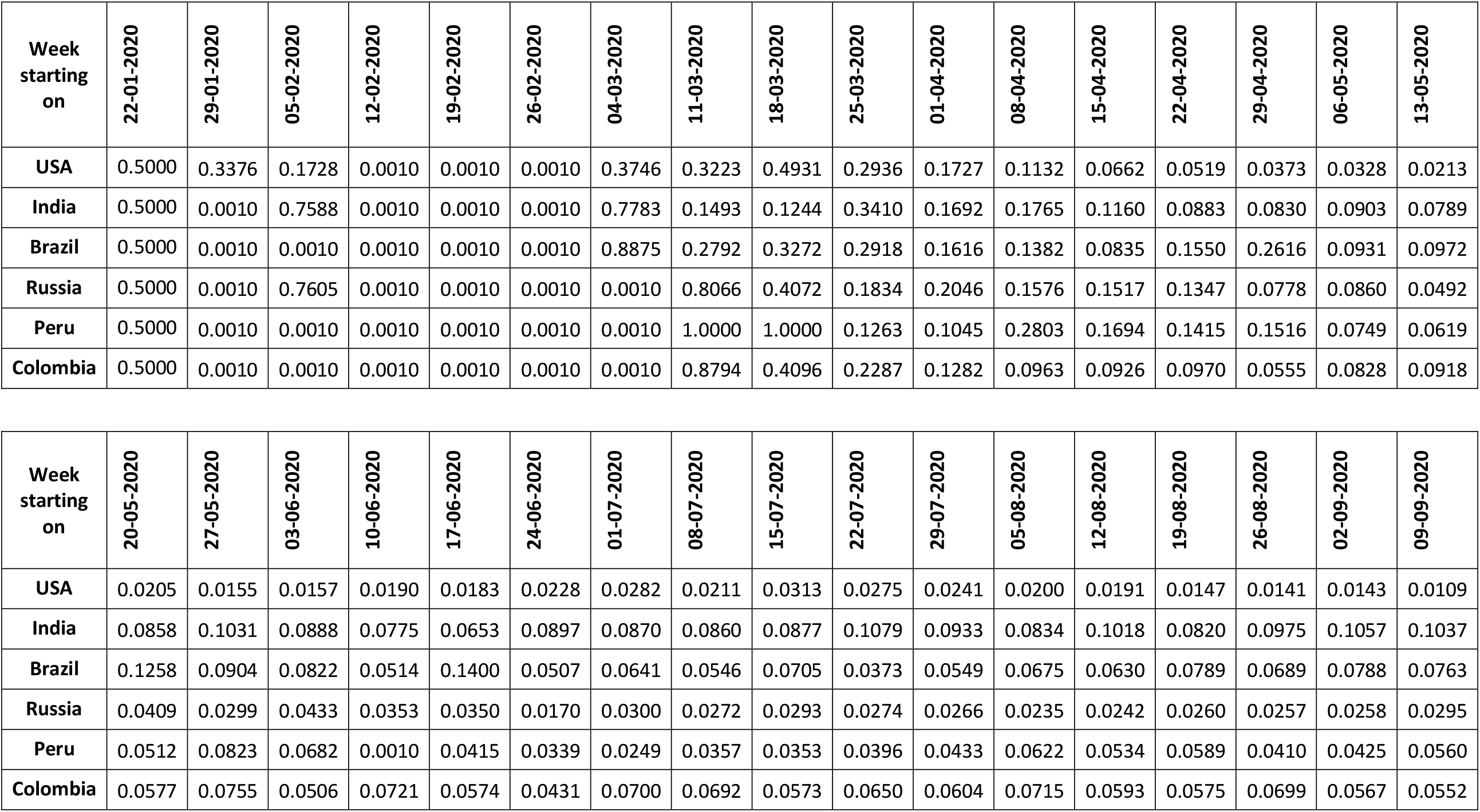
β values for six worst-affected countries

| Week starting on | 22-01-2020 | 29-01-2020 | 05-02-2020 | 12-02-2020 | 19-02-2020 | 26-02-2020 | 04-03-2020 | 11-03-2020 | 18-03-2020 | 25-03-2020 | 01-04-2020 | 08-04-2020 | 15-04-2020 | 22-04-2020 | 29-04-2020 | 06-05-2020 | 13-05-2020 |
| --- | --- | --- | --- | --- | --- | --- | --- | --- | --- | --- | --- | --- | --- | --- | --- | --- | --- |
| USA | 0.5000 | 0.3376 | 0.1728 | 0.0010 | 0.0010 | 0.0010 | 0.3746 | 0.3223 | 0.4931 | 0.2936 | 0.1727 | 0.1132 | 0.0662 | 0.0519 | 0.0373 | 0.0328 | 0.0213 |
| India | 0.5000 | 0.0010 | 0.7588 | 0.0010 | 0.0010 | 0.0010 | 0.7783 | 0.1493 | 0.1244 | 0.3410 | 0.1692 | 0.1765 | 0.1160 | 0.0883 | 0.0830 | 0.0903 | 0.0789 |
| Brazil | 0.5000 | 0.0010 | 0.0010 | 0.0010 | 0.0010 | 0.0010 | 0.8875 | 0.2792 | 0.3272 | 0.2918 | 0.1616 | 0.1382 | 0.0835 | 0.1550 | 0.2616 | 0.0931 | 0.0972 |
| Russia | 0.5000 | 0.0010 | 0.7605 | 0.0010 | 0.0010 | 0.0010 | 0.0010 | 0.8066 | 0.4072 | 0.1834 | 0.2046 | 0.1576 | 0.1517 | 0.1347 | 0.0778 | 0.0860 | 0.0492 |
| Peru | 0.5000 | 0.0010 | 0.0010 | 0.0010 | 0.0010 | 0.0010 | 0.0010 | 1.0000 | 1.0000 | 0.1263 | 0.1045 | 0.2803 | 0.1694 | 0.1415 | 0.1516 | 0.0749 | 0.0619 |
| Colombia | 0.5000 | 0.0010 | 0.0010 | 0.0010 | 0.0010 | 0.0010 | 0.0010 | 0.8794 | 0.4096 | 0.2287 | 0.1282 | 0.0963 | 0.0926 | 0.0970 | 0.0555 | 0.0828 | 0.0918 |

| Week starting on | 20-05-2020 | 27-05-2020 | 03-06-2020 | 10-06-2020 | 17-06-2020 | 24-06-2020 | 01-07-2020 | 08-07-2020 | 15-07-2020 | 22-07-2020 | 29-07-2020 | 05-08-2020 | 12-08-2020 | 19-08-2020 | 26-08-2020 | 02-09-2020 | 09-09-2020 |
| --- | --- | --- | --- | --- | --- | --- | --- | --- | --- | --- | --- | --- | --- | --- | --- | --- | --- |
| USA | 0.0205 | 0.0155 | 0.0157 | 0.0190 | 0.0183 | 0.0228 | 0.0282 | 0.0211 | 0.0313 | 0.0275 | 0.0241 | 0.0200 | 0.0191 | 0.0147 | 0.0141 | 0.0143 | 0.0109 |
| India | 0.0858 | 0.1031 | 0.0888 | 0.0775 | 0.0653 | 0.0897 | 0.0870 | 0.0860 | 0.0877 | 0.1079 | 0.0933 | 0.0834 | 0.1018 | 0.0820 | 0.0975 | 0.1057 | 0.1037 |
| Brazil | 0.1258 | 0.0904 | 0.0822 | 0.0514 | 0.1400 | 0.0507 | 0.0641 | 0.0546 | 0.0705 | 0.0373 | 0.0549 | 0.0675 | 0.0630 | 0.0789 | 0.0689 | 0.0788 | 0.0763 |
| Russia | 0.0409 | 0.0299 | 0.0433 | 0.0353 | 0.0350 | 0.0170 | 0.0300 | 0.0272 | 0.0293 | 0.0274 | 0.0266 | 0.0235 | 0.0242 | 0.0260 | 0.0257 | 0.0258 | 0.0295 |
| Peru | 0.0512 | 0.0823 | 0.0682 | 0.0010 | 0.0415 | 0.0339 | 0.0249 | 0.0357 | 0.0353 | 0.0396 | 0.0433 | 0.0622 | 0.0534 | 0.0589 | 0.0410 | 0.0425 | 0.0560 |
| Colombia | 0.0577 | 0.0755 | 0.0506 | 0.0721 | 0.0574 | 0.0431 | 0.0700 | 0.0692 | 0.0573 | 0.0650 | 0.0604 | 0.0715 | 0.0593 | 0.0575 | 0.0699 | 0.0567 | 0.0552 |

The ε value of USA is high which shows that the asymptomatic cases who are infectious are also high. The values of ε for six worst-affected countries is shown in Table 4. As the infection started late in India, the value is low till February but after that, it increased gradually. At the same time, the exposed value shows a high variation because of various restrictions imposed by the government in India. High ε values requires more testing and quarantining of individuals. The value of espilon in all the countries shows a mix trend due to the restrictions and relaxations offered by the governments from time-to-time. For different countries, the *α* values have been shown in Table 5. *α* denotes the onset rate of infection. This value is constantly high in USA as well as in India. This indicates that the exposed population develops infection symptoms very rapidly in both the countries. In Brazil, Russia, Peru and Colombia, this value shows a mixed trend, depicting a variation in the onset rate. Therefore, the onset period of the appearance of symptoms is almost the same in case of these four nations. The *γ* value represents the Recovery Rate which is high in India and Brazil as shown in Table 6. However, in USA, the *γ* value is lowest, thus indicating a low Recovery Rate and high number of Active Infected cases as compared to other nations. Russia had a high *γ* value in the initial phase of the pandemic. However, this values started decreasing over time. Mu indicates the rate at which infected individuals succumb to the infection. The values of mu for different countries are shown in Table 7. Mu is highest in USA, indicating a large number of fatalities in the country. The fatality rate in India is very low and the same has been depicted by Table 7. Similarly, Russia also shows a very low mu value, and hence a low fatality rate.

**Table 4:**
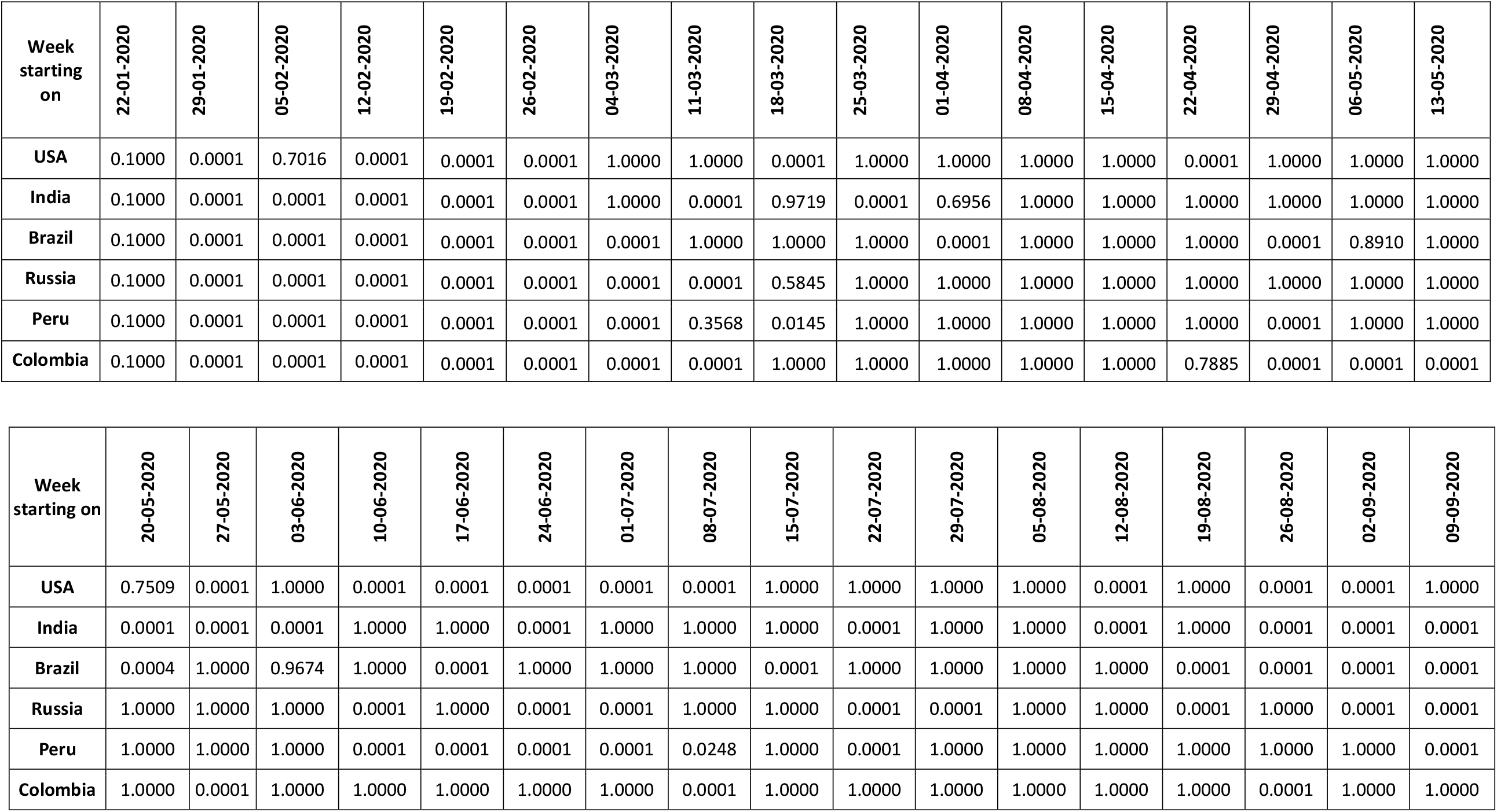
ε values for six worst-affected countries

**Table 5:** *α* values for six worst-affected countries

| Week starting on | 22-01-2020 | 29-01-2020 | 05-02-2020 | 12-02-2020 | 19-02-2020 | 26-02-2020 | 04-03-2020 | 11-03-2020 | 18-03-2020 | 25-03-2020 | 01-04-2020 | 08-04-2020 | 15-04-2020 | 22-04-2020 | 29-04-2020 | 06-05-2020 | 13-05-2020 |
| --- | --- | --- | --- | --- | --- | --- | --- | --- | --- | --- | --- | --- | --- | --- | --- | --- | --- |
| USA | 0.2000 | 1.0000 | 0.1667 | 0.1667 | 0.1667 | 0.1667 | 0.1667 | 0.3117 | 0.4000 | 0.8261 | 0.9824 | 1.0000 | 0.9043 | 0.6460 | 0.5911 | 0.4427 | 0.4711 |
| India | 0.2000 | 0.1667 | 1.0000 | 0.1667 | 0.1667 | 0.1667 | 0.1667 | 0.1852 | 0.3446 | 0.4228 | 0.2834 | 0.4747 | 0.4895 | 0.4969 | 0.5000 | 0.3887 | 0.4566 |
| Brazil | 0.2000 | 0.1667 | 0.1667 | 0.1667 | 0.1667 | 0.1667 | 1.0000 | 0.8418 | 1.0000 | 1.0000 | 0.6525 | 1.0000 | 1.0000 | 0.4390 | 0.2588 | 0.2644 | 0.3719 |
| Russia | 0.2000 | 0.1667 | 1.0000 | 0.1667 | 0.1667 | 0.1667 | 0.1667 | 1.0000 | 0.1667 | 0.1959 | 0.2427 | 0.2735 | 0.2862 | 0.3057 | 0.2568 | 0.2236 | 0.2393 |
| Peru | 0.2000 | 0.1667 | 0.1667 | 0.1667 | 0.1667 | 0.1667 | 0.1667 | 1.0000 | 0.2046 | 0.3294 | 0.4045 | 0.2462 | 0.3480 | 0.1819 | 0.1667 | 0.2225 | 0.2404 |
| Colombia | 0.2000 | 0.1667 | 0.1667 | 0.1667 | 0.1667 | 0.1667 | 0.1667 | 1.0000 | 1.0000 | 0.7709 | 0.7480 | 1.0000 | 1.0000 | 0.1667 | 0.1667 | 0.2385 | 0.2412 |

| Week starting on | 20-05-2020 | 27-05-2020 | 03-06-2020 | 10-06-2020 | 17-06-2020 | 24-06-2020 | 01-07-2020 | 08-07-2020 | 15-07-2020 | 22-07-2020 | 29-07-2020 | 05-08-2020 | 12-08-2020 | 19-08-2020 | 26-08-2020 | 02-09-2020 | 09-09-2020 |
| --- | --- | --- | --- | --- | --- | --- | --- | --- | --- | --- | --- | --- | --- | --- | --- | --- | --- |
| USA | 0.4460 | 0.4882 | 0.8170 | 0.6043 | 0.4516 | 0.3888 | 0.3875 | 0.4303 | 0.6906 | 0.6513 | 0.6134 | 0.5792 | 0.4081 | 0.3332 | 0.2469 | 0.2033 | 0.1667 |
| India | 0.4541 | 0.5339 | 0.4908 | 0.5977 | 0.6698 | 0.7294 | 0.7039 | 0.6269 | 0.5775 | 0.5716 | 0.5972 | 0.6576 | 0.6540 | 0.6452 | 0.6406 | 0.6074 | 0.5879 |
| Brazil | 0.3614 | 0.3669 | 0.3369 | 0.4994 | 0.2707 | 0.1667 | 0.2094 | 0.1667 | 0.1667 | 0.1683 | 0.3352 | 0.4568 | 0.4803 | 0.5126 | 0.4014 | 0.3996 | 0.3677 |
| Russia | 0.2551 | 0.2418 | 0.2134 | 0.1667 | 0.1667 | 0.1667 | 0.2235 | 0.2000 | 0.1788 | 0.1667 | 0.1667 | 0.1677 | 0.1667 | 0.1667 | 0.1667 | 0.1667 | 0.1886 |
| Peru | 0.2748 | 0.2037 | 0.1667 | 0.1667 | 0.3450 | 0.3276 | 0.4488 | 0.6163 | 0.5269 | 0.5798 | 0.6215 | 0.6025 | 0.6320 | 0.3883 | 0.4451 | 0.5951 | 0.3231 |
| Colombia | 0.2313 | 0.2062 | 0.2694 | 0.2394 | 0.1667 | 0.2844 | 0.4343 | 0.3289 | 0.3507 | 0.4430 | 0.4807 | 0.5633 | 0.5293 | 0.7750 | 0.8325 | 0.7727 | 0.9761 |

**Table 6:** *γ* values for six worst-affected countries

| Week starting on | 22-01-2020 | 29-01-2020 | 05-02-2020 | 12-02-2020 | 19-02-2020 | 26-02-2020 | 04-03-2020 | 11-03-2020 | 18-03-2020 | 25-03-2020 | 01-04-2020 | 08-04-2020 | 15-04-2020 | 22-04-2020 | 29-04-2020 | 06-05-2020 | 13-05-2020 |
| --- | --- | --- | --- | --- | --- | --- | --- | --- | --- | --- | --- | --- | --- | --- | --- | --- | --- |
| USA | 0.1000 | 0.0067 | 0.0067 | 0.0428 | 0.0145 | 0.0151 | 0.0167 | 0.0067 | 0.0067 | 0.0067 | 0.0067 | 0.0080 | 0.0070 | 0.0083 | 0.0069 | 0.0133 | 0.0067 |
| India | 0.1000 | 0.1625 | 0.0067 | 0.0067 | 0.1009 | 0.3487 | 0.0067 | 0.0067 | 0.0103 | 0.0142 | 0.0145 | 0.0151 | 0.0196 | 0.0243 | 0.0326 | 0.0301 | 0.0368 |
| Brazil | 0.1000 | 0.1625 | 0.0067 | 0.0067 | 0.0067 | 0.0067 | 0.0067 | 0.0067 | 0.0067 | 0.0067 | 0.0067 | 0.0067 | 0.0095 | 0.1907 | 0.0501 | 0.0443 | 0.0466 |
| Russia | 0.1000 | 0.1625 | 0.0067 | 0.0392 | 0.2280 | 0.0067 | 0.0067 | 0.0067 | 0.0158 | 0.0067 | 0.0103 | 0.0145 | 0.0133 | 0.0114 | 0.0090 | 0.0156 | 0.0182 |
| Peru | 0.1000 | 0.1625 | 0.0067 | 0.0067 | 0.0067 | 0.0067 | 0.0067 | 0.0067 | 0.0067 | 0.0067 | 0.0256 | 0.1702 | 0.0412 | 0.1062 | 0.0067 | 0.0325 | 0.0302 |
| Colombia | 0.1000 | 0.1625 | 0.0067 | 0.0067 | 0.0067 | 0.0067 | 0.0067 | 0.0067 | 0.0067 | 0.0067 | 0.0067 | 0.0083 | 0.0143 | 0.0278 | 0.0154 | 0.0189 | 0.0206 |

| Week starting on | 20-05-2020 | 27-05-2020 | 03-06-2020 | 10-06-2020 | 17-06-2020 | 24-06-2020 | 01-07-2020 | 08-07-2020 | 15-07-2020 | 22-07-2020 | 29-07-2020 | 05-08-2020 | 12-08-2020 | 19-08-2020 | 26-08-2020 | 02-09-2020 | 09-09-2020 |
| --- | --- | --- | --- | --- | --- | --- | --- | --- | --- | --- | --- | --- | --- | --- | --- | --- | --- |
| USA | 0.0067 | 0.0105 | 0.0085 | 0.0082 | 0.0067 | 0.0067 | 0.0067 | 0.0135 | 0.0092 | 0.0074 | 0.0091 | 0.0088 | 0.0082 | 0.0076 | 0.0067 | 0.0067 | 0.0067 |
| India | 0.0447 | 0.0476 | 0.0551 | 0.0417 | 0.0479 | 0.0610 | 0.0644 | 0.0639 | 0.0672 | 0.0616 | 0.0723 | 0.0717 | 0.0840 | 0.0842 | 0.0875 | 0.0838 | 0.0821 |
| Brazil | 0.0387 | 0.0487 | 0.0335 | 0.0519 | 0.0730 | 0.0327 | 0.0494 | 0.0828 | 0.0624 | 0.0555 | 0.0877 | 0.0697 | 0.0712 | 0.0789 | 0.0711 | 0.0822 | 0.0817 |
| Russia | 0.0253 | 0.0307 | 0.0385 | 0.0316 | 0.0333 | 0.0349 | 0.0369 | 0.0324 | 0.0334 | 0.0335 | 0.0348 | 0.0370 | 0.0337 | 0.0313 | 0.0316 | 0.0308 | 0.0288 |
| Peru | 0.0234 | 0.0497 | 0.0273 | 0.0281 | 0.0415 | 0.0374 | 0.0316 | 0.0393 | 0.0339 | 0.0352 | 0.0388 | 0.0355 | 0.0342 | 0.0457 | 0.0292 | 0.0476 | 0.0641 |
| Colombia | 0.0150 | 0.0136 | 0.0227 | 0.0525 | 0.0173 | 0.0307 | 0.0357 | 0.0223 | 0.0272 | 0.0469 | 0.0466 | 0.0482 | 0.0430 | 0.0663 | 0.0800 | 0.0742 | 0.0745 |

**Table 7:** Mu values for six worst-affected countries

| Week starting on | 22-01-2020 | 29-01-2020 | 05-02-2020 | 12-02-2020 | 19-02-2020 | 26-02-2020 | 04-03-2020 | 11-03-2020 | 18-03-2020 | 25-03-2020 | 01-04-2020 | 08-04-2020 | 15-04-2020 | 22-04-2020 | 29-04-2020 | 06-05-2020 | 13-05-2020 |
| --- | --- | --- | --- | --- | --- | --- | --- | --- | --- | --- | --- | --- | --- | --- | --- | --- | --- |
| USA | 0.0100 | 0.0001 | 0.0003 | 0.0230 | 0.0001 | 0.0001 | 0.0223 | 0.0138 | 0.0055 | 0.0055 | 0.0054 | 0.0060 | 0.0052 | 0.0036 | 0.0029 | 0.0021 | 0.0017 |
| India | 0.0100 | 0.1625 | 0.0001 | 0.0001 | 0.0041 | 0.0416 | 0.0001 | 0.0001 | 0.0030 | 0.0042 | 0.0049 | 0.0059 | 0.0053 | 0.0028 | 0.0027 | 0.0033 | 0.0030 |
| Brazil | 0.0100 | 0.1625 | 0.0001 | 0.0001 | 0.0001 | 0.0001 | 0.0001 | 0.0001 | 0.0001 | 0.0055 | 0.0061 | 0.0075 | 0.0063 | 0.0113 | 0.0134 | 0.0093 | 0.0081 |
| Russia | 0.0100 | 0.1625 | 0.0001 | 0.0227 | 0.0072 | 0.0001 | 0.0001 | 0.0001 | 0.0001 | 0.0003 | 0.0015 | 0.0017 | 0.0012 | 0.0013 | 0.0009 | 0.0008 | 0.0006 |
| Peru | 0.0100 | 0.1625 | 0.0001 | 0.0001 | 0.0001 | 0.0001 | 0.0001 | 0.0001 | 0.0001 | 0.0033 | 0.0042 | 0.0111 | 0.0048 | 0.0036 | 0.0045 | 0.0027 | 0.0023 |
| Colombia | 0.0100 | 0.1625 | 0.0001 | 0.0001 | 0.0001 | 0.0001 | 0.0001 | 0.0001 | 0.0001 | 0.0018 | 0.0027 | 0.0038 | 0.0048 | 0.0045 | 0.0031 | 0.0023 | 0.0027 |

| Week starting on | 20-05-2020 | 27-05-2020 | 03-06-2020 | 10-06-2020 | 17-06-2020 | 24-06-2020 | 01-07-2020 | 08-07-2020 | 15-07-2020 | 22-07-2020 | 29-07-2020 | 05-08-2020 | 12-08-2020 | 19-08-2020 | 26-08-2020 | 02-09-2020 | 09-09-2020 |
| --- | --- | --- | --- | --- | --- | --- | --- | --- | --- | --- | --- | --- | --- | --- | --- | --- | --- |
| USA | 0.0012 | 0.0010 | 0.0007 | 0.0007 | 0.0005 | 0.0004 | 0.0003 | 0.0003 | 0.0004 | 0.0003 | 0.0004 | 0.0004 | 0.0003 | 0.0003 | 0.0003 | 0.0002 | 0.0002 |
| India | 0.0022 | 0.0022 | 0.0023 | 0.0024 | 0.0027 | 0.0037 | 0.0016 | 0.0021 | 0.0018 | 0.0018 | 0.0017 | 0.0014 | 0.0015 | 0.0014 | 0.0014 | 0.0013 | 0.0012 |
| Brazil | 0.0067 | 0.0056 | 0.0040 | 0.0031 | 0.0030 | 0.0026 | 0.0019 | 0.0021 | 0.0020 | 0.0018 | 0.0020 | 0.0018 | 0.0017 | 0.0017 | 0.0018 | 0.0016 | 0.0016 |
| Russia | 0.0005 | 0.0006 | 0.0008 | 0.0008 | 0.0007 | 0.0008 | 0.0003 | 0.0009 | 0.0007 | 0.0008 | 0.0005 | 0.0007 | 0.0007 | 0.0005 | 0.0006 | 0.0007 | 0.0004 |
| Peru | 0.0020 | 0.0020 | 0.0021 | 0.0023 | 0.0001 | 0.0024 | 0.0015 | 0.0019 | 0.0018 | 0.0019 | 0.0083 | 0.0005 | 0.0022 | 0.0053 | 0.0011 | 0.0011 | 0.0010 |
| Colombia | 0.0015 | 0.0017 | 0.0016 | 0.0022 | 0.0025 | 0.0019 | 0.0030 | 0.0023 | 0.0023 | 0.0022 | 0.0024 | 0.0025 | 0.0020 | 0.0019 | 0.0020 | 0.0021 | 0.0022 |

## 6. Conclusion

With the rise of COVID-19 cases worldwide, countries need to equip themselves to tackle this pandemic. This can be achieved through accurate predictions of infections in order to help the national governments to prepare policies to control the spread of the infection. These predictions also help in planning both short-term and long-term guidelines to handle the pandemic situation. In this paper, a Modified SEIRD (Susceptible-Exposed-Infected-Recovered-Deceased) model was used to perform COVID-19 predictions for six countries having the highest number of total cases. These include USA, India, Brazil, Russia, Peru and Colombia. This model was chosen as it includes a component which handles infectious asymptomatic individuals and in COVID-19, infections are spread even by asymptomatic persons who are not even aware that they are infected. To predict the trend of the pandemic, epidemiological data up to 9^th^ September 2020 was utilised for experimentation. Short-term predictions were obtained till 31^st^ December 2020, while long-term predictions were performed till the end of year 2023. For short-term predictions, the results were statistically analysed by using t-test with 99% confidence. As per predictions by Modified SEIRD model, it was found that the peak of COVID-19 in USA is expected around July 2022 with 2,29,457 daily confirmed cases, while in India, it is expected around May 2021 with 21,26,406 cases coming daily. It was also observed that Russia and Brazil witnessed the peak of infection in May 2020 and July 2020 respectively, whereas for Peru and Colombia the peak of COVID-19 infection was witnessed in August 2020.

## Data Availability

Open data has been used for implementation

https://www.covid19india.org/

## Notes

### Competing Interest Statement

The authors have declared no competing interest.

### Funding Statement

No funding

### Author Declarations

No such approvals are required

